# Enhancer RNA transcriptome-wide association study reveals an atlas of pan-cancer susceptibility eRNAs

**DOI:** 10.1101/2024.02.29.24303580

**Authors:** Wenyan Chen, Zeyang Wang, Jianxiang Lin, Shuxin Chen, Hui Chen, Xuelian Ma, Xudong Zou, Xing Li, Yinuo Wang, Yangmei Qin, Xixian Ma, Yunbo Qiao, Lei Li

## Abstract

Many cancer risk variants are located within enhancer regions and lack sufficient molecular interpretation. Here, we constructed the first comprehensive atlas of enhancer RNA (eRNA)- mediated genetic effects from 28,033 RNA sequencing samples across 11,606 individuals, identifying 11,757 eRNA quantitative trait loci (eRNA-QTLs) significantly associated with eRNA expression. Mechanistically, eRNA-QTLs frequently altered binding motifs of transcription factors. In addition, 28.48% of cancer risk variants were strongly colocalized with eRNA-QTLs. We further performed an eRNA-based transcriptome-wide association study and identified 626 cancer susceptibility eRNAs across 23 cancer types. 54.9% of the eRNA target genes were overlooked by traditional gene expression studies, and most are essential for cancer cell proliferation. To substantiate our findings, we confirmed the enhancer functionality of two newly identified susceptibility eRNAs, *CCND1e* and *SNAPC1e*, through CRISPR-based inhibition, resulting in a marked decrease in the expression of their respective target genes, consequently suppressing the proliferation of prostate cancer cells. Our study underscores the essential role of eRNA in unveiling new cancer susceptibility genes and establishes a strong framework for enhancing our understanding of human cancer etiology.

## Introduction

Genome-wide association studies (GWASs) have identified numerous single-nucleotide polymorphisms (SNPs) associated with complex human traits and disorders ^1^. Most are located in noncoding regions of the genome ^2^, particularly enhancer regions ^3–5^. Expression quantitative trait loci (eQTLs) often act as a crucial link between GWAS SNPs and disease phenotypes. Although eQTLs and other molecular QTLs ^6^ provide valuable insights into regulation of nearby gene transcription, the functional roles of these disease-associated genetic variants spanning noncoding RNAs remain largely unknown.

Enhancer RNAs (eRNAs) are a subclass of noncoding RNAs transcribed from active enhancer regions within the genome ^7–9^. eRNA transcription is closely associated with RNAPII binding and epigenetic modifications, such as histone marks H3K27ac and H3K4me1 ^10^. Transcribed eRNAs can act as independent regulators that modulate the expression of nearby genes. For instance, p53-bound enhancer regions can generate eRNAs that stimulate their transcription in response to DNA damage, resulting in activation of p53-dependent cell cycle arrest and apoptosis ^11,12^. In addition, eRNAs make major contributions to the modulation of disease progression and trait development ^13^, especially in human cancers ^14,15^. Specific examples include *CCAT1e*, an eRNA transcribed from the *CCAT1* locus, which interacts with the transcription factor (TF) TCF7L2 to activate the wnt signaling pathway, thereby promoting colon cancer cell proliferation and invasion ^14,16,17^. Additionally, *KLK3e* and *PSAe* are eRNAs transcribed from enhancers regulated by the androgen receptor that affect androgen-induced gene activation and potentially contribute to the development of prostate cancer ^18,19^. In addition, eRNAs play an important role in the pathology of neurodegenerative disorders ^20,21^, contributing to the loss of neuronal function and viability by regulating genes critical for synaptic plasticity, neuroinflammation, and protein misfolding. However, despite these individual examples, the prevalence and magnitude of eRNAs influencing cancer susceptibility remain largely unknown.

Several specialized experimental profiling methods, such as nuclear run-on followed by cap-selection assay (GRO/PRO-cap) and self-transcribing active regulatory region sequencing (STARR-seq), can identify actively transcribed eRNAs ^22^. However, these approaches have not been widely adopted by population-scale studies. By contrast, RNA sequencing (RNA-seq) has been extensively used in many genomic projects, such as Genotype-Tissue Expression (GTEx)^23^. In addition, recent studies demonstrate the ability of RNA-seq to identify eRNAs ^15^. Despite extensive attention to eRNAs, however, their genetic impact and role in cancer susceptibility remains poorly understood.

In this study, we performed the first large-scale, systematic analysis assessing eRNA-mediated genetic effects on 49 human normal tissues and 31 tumor types by analyzing an extensive dataset of 28,033 RNA-seq samples from 11,606 individuals. eRNA-QTLs often disrupt the binding motifs of TFs, leading to altered expression of corresponding eRNAs. We validated this using CRISPR-based base editing experiments. Additionally, we conducted eRNA expression transcriptome-wide association studies (eRNA-TWAS) that facilitated the functional characterization of cancer risk loci. We also validated the enhancer activity of two newly identified susceptibility eRNAs (*CCND1e*, *SNAPC1e*) in regulating their target expression by CRISPR-based inhibition, consequently suppressing the proliferation of prostate cancer cells. Lastly, we developed a comprehensive data portal called the eRNA-QTL atlas (http://bioinfo.szbl.ac.cn/eRNA-QTL-atlas/), which provides a valuable resource for the research community by granting access to the extensive information generated in this study. Overall, our findings substantially advance our understanding of the eRNA-mediated genetic effects contributing to cancer risk.

## Results

### Atlas of eRNA-mediated genetic effects

To systematically detect eRNAs in primary human tissues, we annotated and quantified the expression of eRNA based on RNA-seq data following previously described methods ^10^. We identified 12,509 expressed eRNAs from 17,265 RNA-seq samples across 49 human normal tissues using the GTEx v8 dataset. We also included 9,111 expressed eRNAs from 10,768 RNA-seq samples across 31 human tumor types using The Cancer Genome Atlas (TCGA) dataset (Fig. 1A). To assess whether eRNA expression profiles effectively differentiate human tissues/organs, we performed clustering analysis of eRNA expression across all samples. This analysis unveiled distinct patterns of eRNA expression among various human tissues. eRNA expression originating from the same organ tended to cluster together and was discernibly different from expression in other organs, as exemplified by distinctions observed between the artery and heart (Fig. S1).

**Fig. 1.**
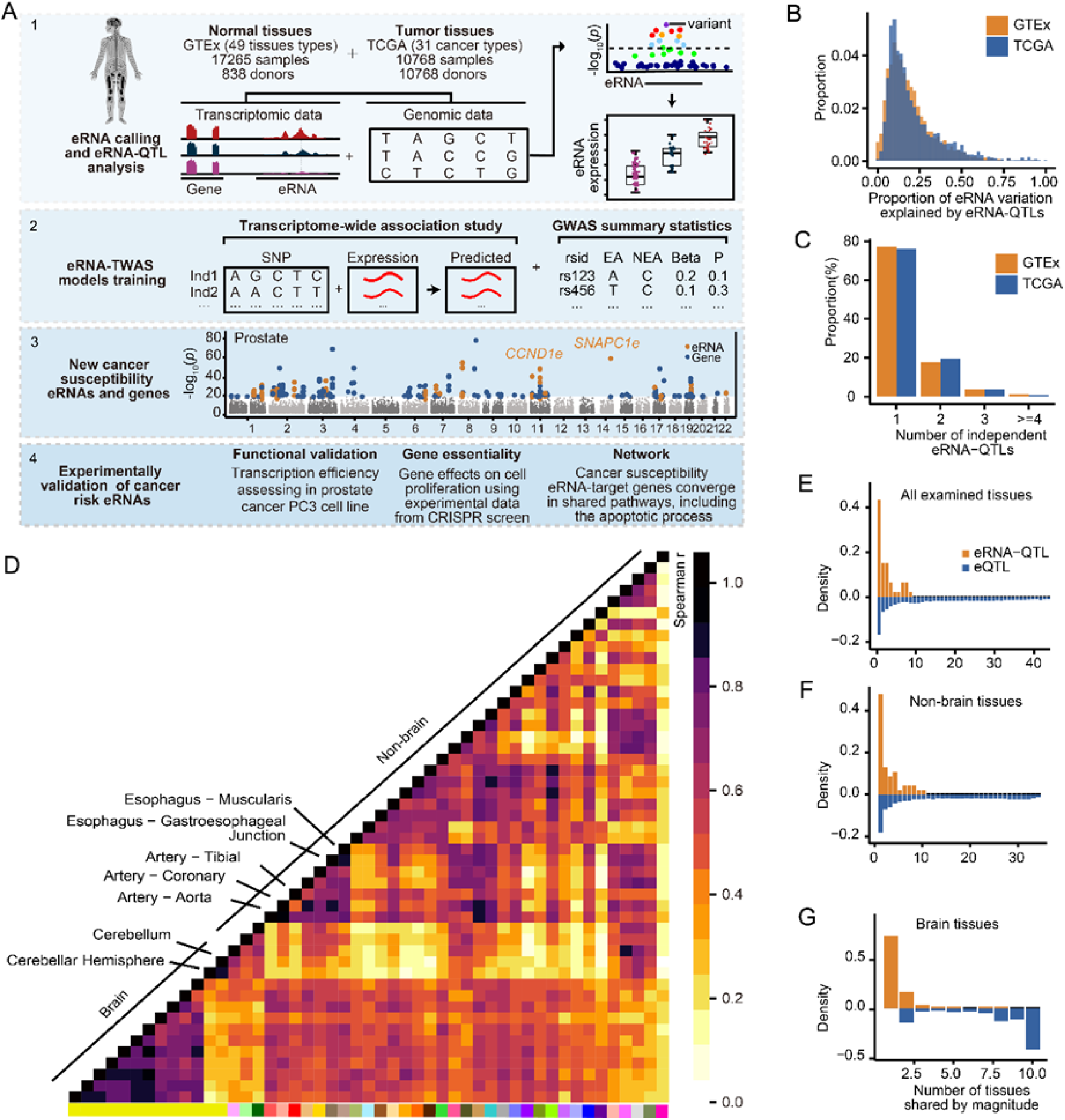
QTL mapping on eRNA transcriptome profiles across 48 tissues. (A) Overview of the study and data. We integrated RNA-seq and genotype data from GTEx and TCGA to develop a reference panel for eRNA-QTL analysis and eRNA-TWAS modeling. We then employed eRNA-TWAS analysis to identify cancer susceptibility eRNA-link genes using cancer GWAS summary statistics and eRNA-TWAS models. The biological functions of these cancer susceptibility eRNA-link genes were further validated by data analysis and experimental approach. (B) Average fraction of eRNA variations that could be explained by eRNA-QTLs. The y-axis represents the proportion of eRNAs across all human normal (GTEx) tissues and tumor (TCGA) types studied. (C) Proportion of independent eRNA-QTLs across all human normal (GTEx) tissues and tumor (TCGA) types. (D) Pairwise eRNA-QTL sharing by magnitude among tissues. eRNA-QTL sharing patterns were assessed pairwise across various tissues by examining the Spearman correlation between mashR effect sizes for each tissue pair. The results are displayed in matrix format, with each cell representing the correlation value for a particular tissue pair. To identify shared eRNA-QTLs between two tissues, the top eRNA-QTLs that attained significance (local false sign rate < 0.05) in at least one of the two tissues were selected. The proportion of shared eRNA-QTLs was then plotted, whereby only those with effect estimates of the same sign and within a factor of 2 in size were included. The hierarchical clustering algorithm was utilized to arrange the tissues based on their similarity in eRNA-QTL sharing patterns. (E) Proportion of tissues sharing lead eRNA-QTLs/eQTLs across all 49 examined tissues, among non-brain tissues (F) and brain tissues (G).

To investigate the impact of genetic variations on eRNA expression, we identified 11,757 eRNA-QTLs (i.e., genetic variants associated with eRNA expression) associated with 89.75% (12,509/13,937) of annotated eRNAs using QTLtools ^24^ (Fig. 1A). Notably, genomic inflation lambdas for eRNA-QTLs ranged from 0.95 to 1.17 across all tissues, indicating that population stratification was tightly controlled. We found that the number of eRNA-QTLs varied across 49 normal tissues from GTEx, ranging from 57 eRNA-QTLs associated with 57 eRNAs in the brain substantia nigra to 3,022 eRNA-QTLs associated with 2,533 eRNAs in the testis (Table S1). Interestingly, we observed that eRNA-QTLs were more prevalent in tissues with larger sample sizes (Rho = 0.99, *P* < 0.001; Table S1). In addition, we identified 9,316 eRNA-QTLs across 31 tumor types from TCGA. Several previously known eRNA-QTLs were successfully recovered, such as the eRNA-QTL rs72700813 (Fig. S2A) linked to the modulation of *GOLPH3L* eRNA (*GOLPH3Le*) expression, which is closely associated with neurological disorders ^25^ (Fig. S2B).

To estimate the heritability of eRNAs, we used genome-wide complex trait analysis (GCTA) ^26^ to quantify the eRNA expression variation that can be explained by eRNA-QTLs. Our analysis revealed that eRNA-QTLs collectively explain an average of 24.4% and 20.97% of eRNA variation in normal tissues and tumor types, respectively (Fig. 1B). For example, *NET1* eRNA (*NET1e*), an oncogene in breast, prostate, and liver cancer ^14^, had a high heritability estimate of 0.46 (*P* < 2.2×10^-16^). Also, *KANSL1* eRNA (*KANSL1e*), which disrupts autophagy and thereby results in memory impairment and neurodegeneration ^20^, had a high heritability estimate of 0.90 (*P* = 6.49×10^-11^). Next, we performed conditional stepwise regression to identify independent genetic variants associated with eRNA expression. In contrast to eGenes (i.e., genes with at least one eQTL), for which up to 50% have at least two independent eQTLs in the given tissue ^23^, our results indicate that 77.25% and 76.03% of eRNAs contained one independent eRNA-QTL in normal tissues and tumor types, respectively (Fig. 1C). For example, *BRCA1* eRNA (*BRCA1e*), located upstream of the *BRCA1* gene, contains an independent eRNA-QTL rs11650272 (*P* = 1.63×10^-10^; Fig. S2B).

To explore the sharing patterns of eRNA-QTLs across tissues, we applied the multiple adaptive shrinkage (Mash) model ^27^ to calculate pairwise eRNA-QTL sharing. Generally, eRNA-QTLs cluster into two distinct groups: brain and non-brain tissues (Fig. 1D). Approximately 73.4% of eQTLs are shared across multiple tissues ^27,28^. By contrast, we found that only 26.67% and 52.17% of eRNA-QTLs were shared among brain and non-brain tissues, respectively (Fig. 1, E to G). This suggests that eRNA-QTLs display a higher degree of tissue specificity than eQTLs. Using these results, we constructed a comprehensive atlas of eRNA-mediated genetic effects across 49 human normal tissues and 31 tumor types, highlighting the discriminative potential of eRNA-QTLs among various biological tissues.

### eRNA-QTLs have distinct molecular features

We further conducted functional annotations to determine the positional distribution of lead eRNA-QTLs, defined as the genetic variants exhibiting the strongest associations with their corresponding eRNAs. As expected, most lead eRNA-QTLs were enriched within eRNA regions (Fig. S2C). By contrast, only 11.75% of lead eRNA-QTLs overlapped with eQTLs (Fig. S2D), demonstrating that eRNA-QTLs may represent unique genetic regulatory information. We further conducted functional annotation enrichment of lead eRNA-QTLs and eQTLs using torus^29^ and found that eRNA-QTLs were significantly enriched in enhancer (4.44-fold enrichment compared with matched background SNPs with the same annotation in the whole genome; two-sided Fisher’s exact test, *P* = 8.43×10^-59^) and promoter (1.8-fold enrichment compared with matched background SNPs with the same annotation in the whole genome; two-sided Fisher’s exact test, *P* = 2.43×10^-114^) regions (Fig. 2A).

**Fig. 2.**
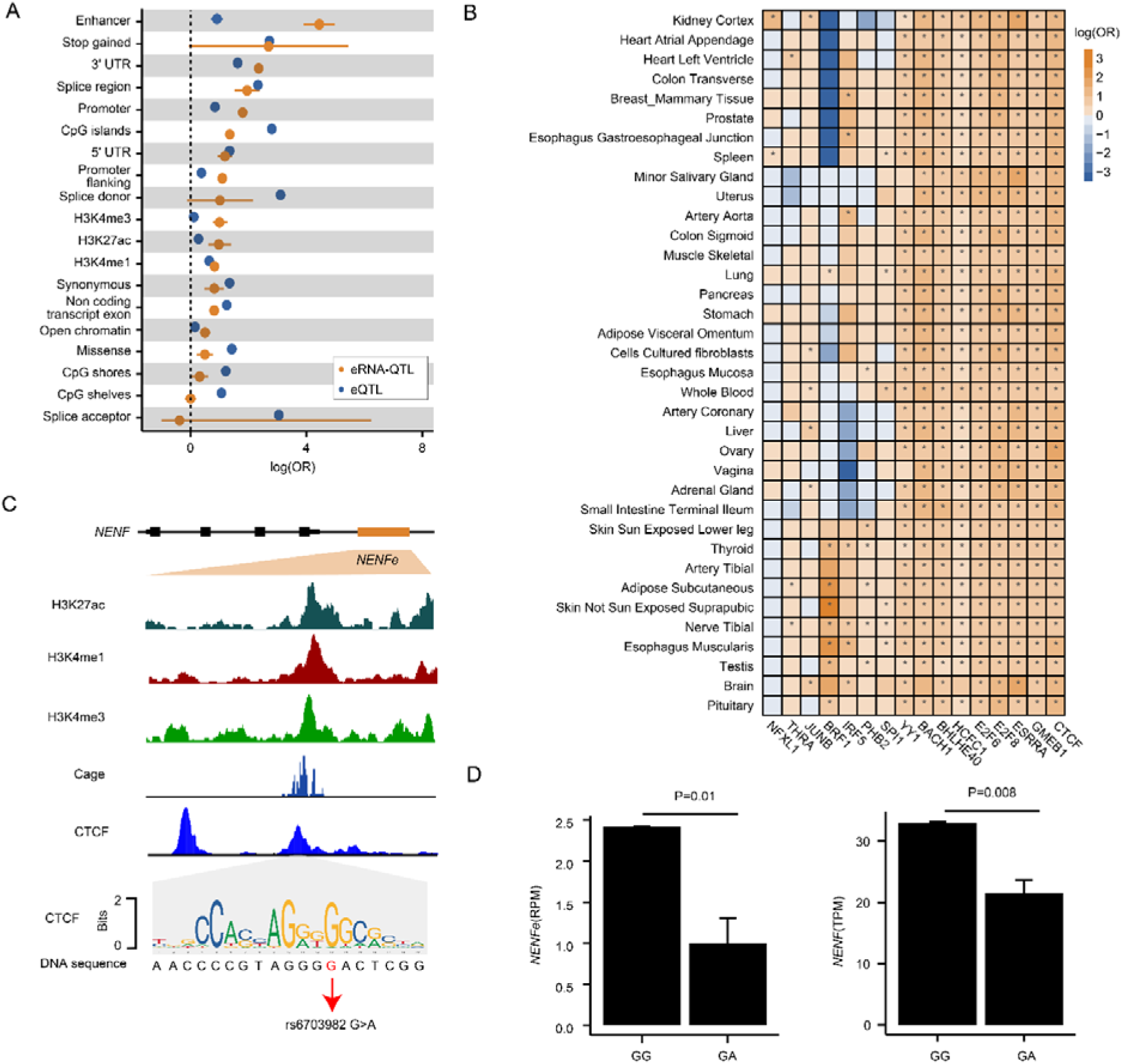
Functional annotation of eRNA-QTLs. (A) Enrichment of eRNA-QTLs and eQTLs for different genome annotations. Each dot in the figure represents the log-transformed odds ratio (OR), and the lines indicate the corresponding 95% confidence interval. Enrichment was assessed using all SNPs through torus based on the eRNA-QTL results obtained using QTLtools and the eQTL results obtained from the GTEx Portal. The peak files of histone modification markers were downloaded from the Roadmap Epigenomics Project. (B) eRNA-QTL were enriched in TFBSs across tissues. The enrichment values correspond to the maximum-likelihood estimated log(OR), and the whiskers on the plot indicate values below the significant FDR threshold of 0.05. (C) The genome browser displays histone modification data, CAGE data, and CTCF binding site data within the *NENF* eRNA (*NENFe*) locus. Histone modification data were downloaded from the Roadmap Epigenomics Project, CAGE data were obtained from FANTOM5, and CTCF binding site data were acquired from Cistrome. A DNA logo is presented representing the CTCF-binding motif based on previously reported consensus CTCF binding sites ^55^. The height of each letter in the logo indicates the relative frequency of occurrence of the corresponding nucleotide at that specific position. (D) The A allele of rs6703982 was observed to downregulate the expression of *NENF* eRNA (*NENFe*) and its target gene *NENF*.

To examine the regulatory mechanisms of eRNA-QTLs and their target genes, we employed a Bayesian network (BN) ^30^ to analyze the extent of causal regulation of eRNA-QTL on target gene expression via eRNA. We evaluated three causality models: a causal model (SNP→eRNA→gene) in which eRNA-QTLs affect eRNA expression and then influence gene expression; a reactive model (SNP→gene→eRNA) in which eRNA-QTLs affect nearby gene expression and then influence eRNA expression; and an independent model (eRNA←SNP→gene) in which eRNA-QTLs affect eRNA expression and nearby gene expression independently (Fig. S3A). We identified 15,879 SNP-eRNA-gene triplets across various tissues, with a median of 375 triplets per tissue. Notably, a median of 48.67% of SNP-eRNA-gene triplets was consistent with a causal model (Fig. S3B). Interestingly, the SNPs in the causal model were more proximate to eRNAs, whereas the SNPs in the reactive model were closer to the target genes (Fig. S3, C and D; two-sided Wilcoxon rank-sum test, *P* < 2.2×10^-16^). To investigate whether these models were enriched in active histone marks associated with gene expression, we utilized FORGE2 ^31^ to map histone mark enrichment for 39 cell types from the Roadmap Epigenomics Project ^32^. The causal and reactive models were enriched with active histone marks associated with active enhancers or transcription, such as H3K4me1 and H3K4me3 (Fig. S3, E and F). Also, all models of SNPs were relatively common in active but not repressive histone marks, suggesting the involvement of eRNAs in regulating transcriptional activation (Fig. S3F). Our findings also reveal that causal model eRNAs were more frequently associated with enhancer-promoter loops than reactive model eRNAs (Fig. S3G, Fisher’s exact test, *P* = 7.17×10^-3^) and independent model eRNAs (Fisher’s exact test, *P* = 1.26×10^-9^), illustrating the intricate interactions contributing to transcriptional regulation. Conversely, reactive model eRNAs were more frequently associated with the chromatin remodeling process than causal model eRNAs (Fig. S3H, Fisher’s exact test, *P* = 0.03) and independent model eRNAs (Fisher’s exact test, *P* = 0.04). Taken together, our findings highlight the causal role of eRNA-QTLs in regulating gene expression.

### eRNA-QTLs disrupt known and novel transcription factor binding sites

To investigate the potential genetic mechanism contributing to eRNA expression, we hypothesized that certain eRNA-QTLs could disrupt transcription factors binding sites (TFBSs), leading to alterations in eRNA expression. We analyzed ChIP-seq data for 194 TFs and 17 histone marks from The Encyclopedia of DNA Elements (ENCODE) project ^33^. 158 TFs were identified that exhibited significant enrichment in eRNA-QTLs in the GTEx dataset, and 171 TFs showed significant enrichment in eRNA-QTLs in the TCGA dataset. Notably, several TFs were enriched in eRNA-QTLs across a diverse set of tumor types. Specifically, *MYC* displayed significant enrichment in eRNA-QTLs across 16 tumor types (Fig. S4A). Several TFs are known to be associated with eRNA regulation. For example, SNP rs17776622, located within the core consensus binding motif of *YY1*, was significantly associated with the expression of *SOX7* eRNA (*SOX7e*) (Fig. S4B). Moreover, *SOX7e* displayed tight co-regulation with *SOX7* (Fig. S4C), suggesting that eRNA-QTL rs17776622 could affect *SOX7* gene expression by altering YY1 binding motifs. Pathway enrichment analysis demonstrated that these TFs are involved in several important biological processes, including transcription activator activity (Fisher’s exact test, adjusted *P* = 9.80**×**10^-7^) and polymerase II transcription activator activity (Fisher’s exact test, adjusted *P* = 1.88**×**10^-6^) (Fig. S4D). Interestingly, we observed significant enrichment of eRNA-QTLs in *CTCF* binding sites across all tissues (Fig. 2B). To validate our hypothesis, we performed CRISPR-based base editing experiments to mimic the A allele of rs6703982, the SNP located at the CTCF binding site (Fig. 2C), in HEK293T cells and successfully obtained two clones containing heterozygous G/A mutations (Fig. S4E). Consistent with our prediction, disrupting rs6703982 inhibited the expression of rs6703982-associated *NENF* eRNA (*NENFe*) and its potential target *NENF* (Fig. 2D). Thus, our findings suggest that a proportion of altered eRNA expression is elicited by eRNA-QTLs that disrupt the binding motifs of TFs.

### eRNA-QTLs strongly contribute to cancer heritability

To investigate the proportion of disease-associated variants in the eRNA region, we analyzed the genomic distribution of disease-associated fine-mapped variants (95% credible sets) from CAUSALdb ^34^. Cancer-associated variants showed the highest enrichment in the eRNA region compared with other disease domains, highlighting the potential role of eRNAs in carcinogenesis (Fig. 3A). For example, the prostate cancer lead SNP rs4857906 (*P* = 1.05**×**10^-9^) was identified in the eRNA region of the *GATA2* gene, which encodes a member of the GATA family of zinc-finger TFs (Fig. 3B). Remarkably, it exhibited the highest posterior probability of being a causal SNP of prostate cancer (Fig. 3B).

**Fig. 3.**
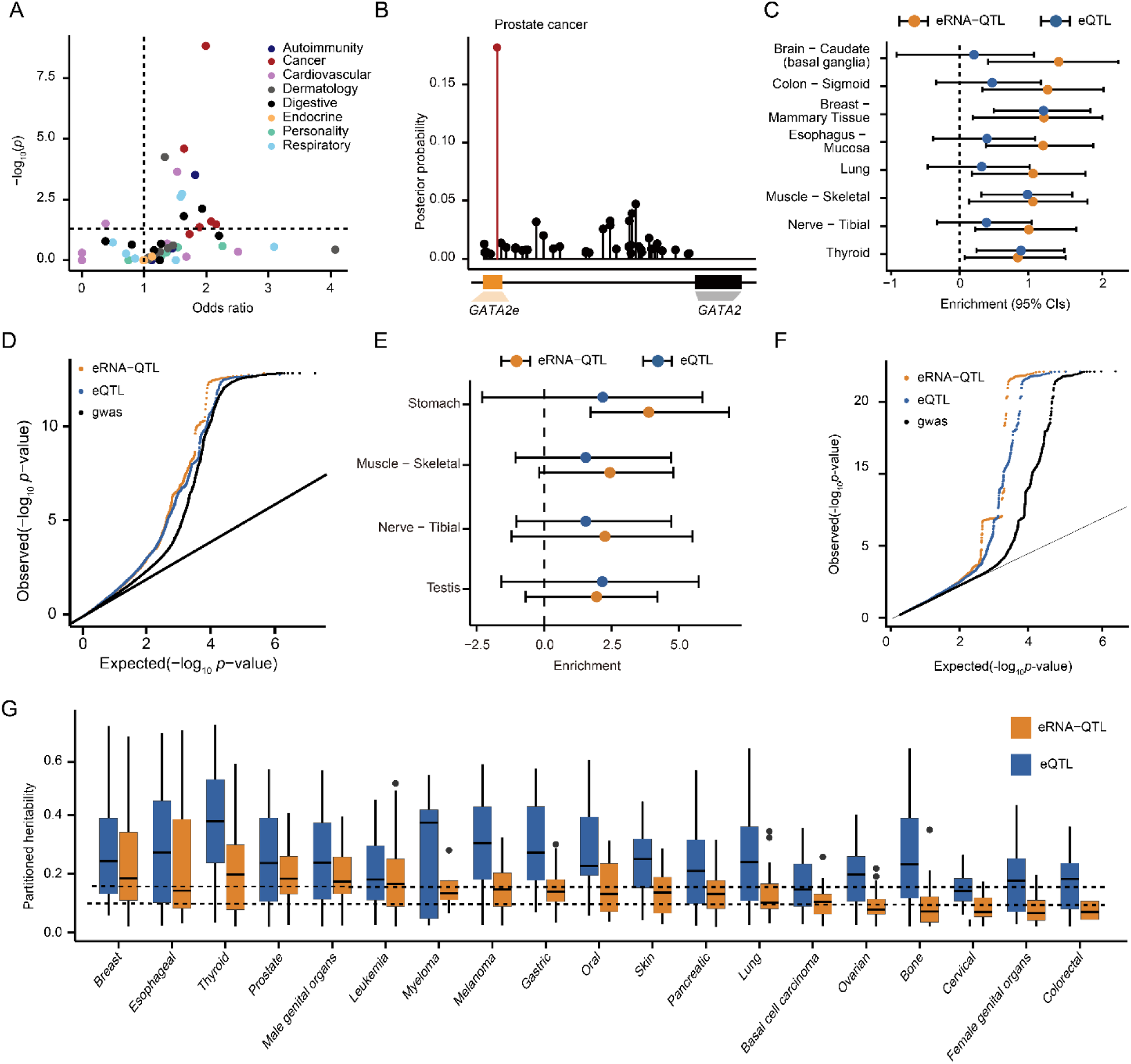
Contribution of eRNA-QTLs to cancer heritability. (A) Cancer causal variants were more significantly enriched in eRNA regions. To investigate the genomic distributions of disease-associated fine-mapped variants (95% credible sets), we conducted an analysis of these variants within eRNA regions across various disease domains sourced from CAUSALdb. (B) The highest PP of a causal variant for prostate cancer was observed in eRNA regions. The *GATA2* eRNA (*GATA2e*) region contained eight causal SNPs associated with prostate cancer. Notably, rs4857906 (red color), an SNP located within the *GATA2e* region, exhibited the highest PP with prostate cancer. (C) Tissues with eRNA-QTL enrichment but no eQTL enrichment for breast cancer. Enrichment values (i.e., effect size) were calculated using functional GWAS quantifying relationships between trait-associated variants and eRNA-QTLs/eQTLs. A positive value indicates that variants with stronger association evidence in GWAS are more likely to be eRNA-QTLs/eQTLs. Estimated lower- and upper-bound 95% confidence intervals for enrichment values are also shown. (D) Example quantile-quantile plot (QQ plot) showing the nominal *P*-values of breast cancer GWAS SNPs, which were primarily annotated by eRNA- QTLs (yellow) and eQTLs (blue). Each dot represents a GWAS SNP. All breast cancer GWAS nominal *P*-values are also shown as controls (black). (E) Tissues with eRNA-QTL enrichment but no eQTL enrichment for gastric cancer. (F) Example QQ plot showing the nominal *P*-values of gastric cancer GWAS SNPs. (G) A significant contribution to the heritability of cancers by eRNA-QTL/eQTL. Cancer types were grouped on the x-axis. For each cancer type, the yellow and blue colors indicate the contribution of eRNA-QTLs and eQTLs to the heritability of cancers, respectively.

To further evaluate the enrichment of eRNA-QTLs on cancer traits, we compiled and curated 57 GWAS summary statistics covering 23 cancer types from the literature (Table S2) and performed functional GWAS analyses ^35^. Among these 395 tissue-trait pairs, 20.76% showed significant associations with eQTLs, and 21.77% showed significant associations with eRNA- QTLs. Among the eRNA-QTLs tissue-trait pairs, 3.54% displayed a more significant effect than eQTLs, and 16.20% were exclusive to eRNA-QTLs. Many eRNA-QTLs were enriched in tissues relevant to their diseased states, such as mammary breast tissue for breast cancer (enrichment = 1.23; Fig. 3C), stomach tissue for gastric cancer (enrichment = 3.89), and uterus tissue for leiomyoma of the uterus (enrichment = 2.75). Another instance was observed in stomach tissue, in which there was enrichment of eRNA-QTL but not eQTL in gastric cancer (Fig. 3, E and F). To investigate the extent of cancer heritability attributed to eRNA-QTLs, we used linkage disequilibrium score regression ^36^ to estimate the proportion of eRNA-QTLs associated with cancer heritability. We observed that a relatively high proportion of heritability, ranging from a mean of 5.41% to 29.61% per trait, could be explained by eRNA-QTLs (Fig. 3G). Altogether, these findings suggest that eRNA-QTLs make strong contributions to cancer heritability.

### Most colocalized cancer loci were exclusively detected by eRNA-QTLs

To identify the subset of eRNAs contributing to cancer risk through genetic effects, we performed summary data-based Mendelian randomization (SMR) ^37^ and colocalization ^38^ analyses to evaluate whether cancer risk loci share the same causal variants as eRNA-QTLs. A total of 85 cancer risk loci were found to colocalize with eRNA-QTLs (Fig. 4A; Table S3 and S4). An average of 28.48% cancer risk loci colocalized with eRNA-QTLs, and 70.02% colocalized cancer loci were exclusively detected by eRNA-QTL but not eQTL (Fig. 4B). For instance, eRNA-QTL rs6866783 in *CLPTM1L* eRNA (*CLPTM1Le*) exhibited strong colocalization with cervical cancer (posterior inclusion probability (PP)_H4_ = 0.92), whereas there was no colocalization with the eQTL of *CLPTM1L* (PP_H4_ = 0.01; Fig. 4C), which encodes cleft lip and palate transmembrane protein 1-like and could increase susceptibility to various cancers. In another instance, eRNA-QTL rs62431527 in *CNR1* eRNA (*CNR1e*) exhibited strong colocalization with ovarian cancer (PP_H4_ = 0.99), whereas there was no colocalization with the eQTL of *CNR1* (PP_H4_ = 0.00; Fig. 4D). Furthermore, genetic variant rs56687477, associated with an increased risk of breast cancer, was correlated with higher expression of *ZNF703* eRNA (*ZNF703e*) (PP_H4_ = 0.99) and *ZNF703* (PP_H4_ = 0.98, Fig. S5A), which encodes zinc finger protein 703, a commonly observed oncogene in luminal B breast cancer.

**Fig. 4.**
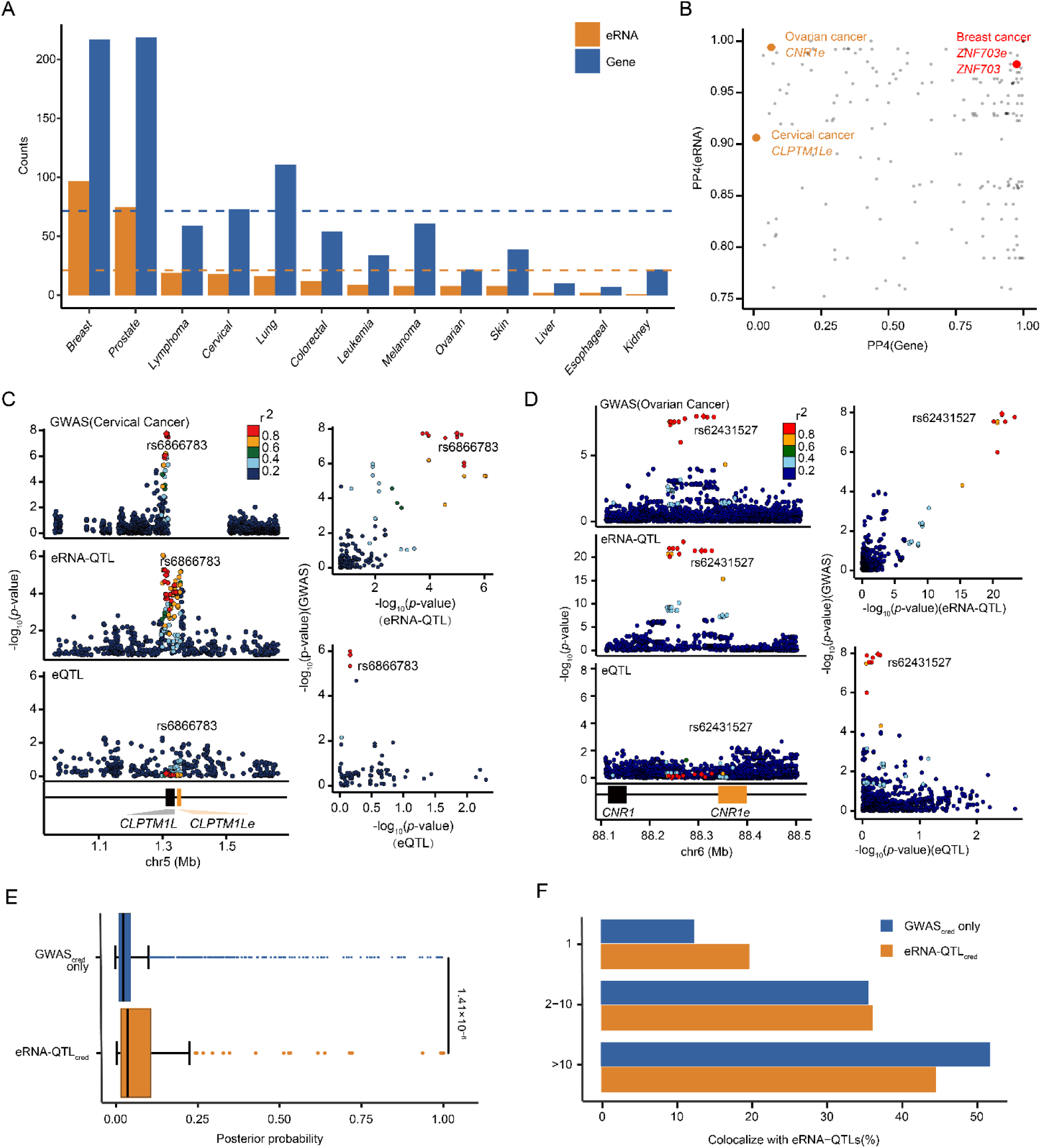
Locus colocalization and SMR analyses. (A) Number of colocalization events for eRNA-QTL and eQTL. Cancer types are grouped on the x-axis. For each cancer type, yellow and blue colors indicate that the colocalization events could be explained by eRNA-QTL or eQTL, respectively. (B) Scatter plot showing colocalization PPs for significant genes and eRNA, respectively. eRNA-QTLs/eQTLs were considered be to colocalized with GWAS results if PP_H4_ exceeded 75% and PP_H4_/(PP_H3_ + PP_H4_) was ≥ 0.9. PP_H3_: independent causal variants; PP_H4_: colocalized causal variants. Examples of colocalization events that could only be explained by eRNA-QTL are marked in yellow, and examples of colocalization events that could be explained by both eRNA-QTL and eQTL are marked in red. (C) Independent eRNA-QTLs depict variants with regulation on the eRNA level but not the mRNA displayed by the significant SNP-eRNA pair rs6866783-*CLPTM1Le*. Variants are represented by points colored relative to LD with the candidate variant rs6866783 (red, ≥ 0.8; orange, 0.6–0.8; green, 0.4–0.6; light blue, 0.2–0.4; dark blue, < 0.2). LD data from 1,000 Genomes (phase 3). (D) Independent eRNA-QTLs depict variants with regulation on the eRNA level but not the mRNA level displayed by the significant SNP-eRNA pair rs62431527-*CNR1e*. (E) Distribution of PP for all fine-mapped GWAS variants (95% credible set in GWAS, GWAS_cred_) that were also fine-mapped eRNA-QTL variants (eRNA-QTL_cred_) vs. GWAS_cred_ variants only. Mann-Whitney *P-*value is shown. (F) Integration of cancer-credible GWAS variants with credible sets from colocalizing eRNA-QTLs increased fine-mapping resolution. The bar plot shows the proportion of independent loci identified as candidate causal variants before and after restricting for QTL variants. The credible set sizes are binned into three groups (1, 2-10, and > 10).

To further investigate whether eRNA-QTLs are enriched at causal cancer risk loci, we conducted fine mapping on colocalized eRNA-QTLs and compared them with cancer risk loci within a 95% credible set. Our analysis revealed that cancer-credible set variants shared with eRNA-QTLs had a significantly higher PP than those not shared with eRNA-QTLs (Mann-Whitney U test, *P* = 1.41×10^-6^; Fig. 4E). Moreover, incorporating eRNA-QTLs significantly improved the genetic resolution of cancer-credible sets, leading to the identification of 19.58% risk loci with one potential causal variant compared with only 12.45% risk loci when eRNA-QTLs were not considered (Fig. 4F). These results suggest that eRNA-QTLs contribute distinguishably to cancer risk variants.

### The landscape of cancer susceptibility eRNAs across 23 cancer types

To systematically identify eRNA-linked susceptibility genes associated with human cancers, we adapted the traditional TWAS methodology (using FUSION software ^39^) by examining the association between eRNA expression and GWAS statistics (referred to as eRNA-TWAS). In each dataset, we employed a mixed-linear model to estimate the heritability of eRNA expression. This estimation was based on *cis*-SNPs located near the eRNA in a reference panel consisting of cohorts with matched RNA-seq and genotype data. Only eRNAs with significant heritability estimates (*cis*-h^2^) at a *P* < 0.05 were included in further analyses. For each FUSION-trained model, such as the best linear unbiased predictor (BLUP), elastic-net regression (ENET), and lasso regression (LASSO), we utilized cross-validation to select the model that offered the most accurate prediction for eRNA-TWAS for each specific eRNA. In total, we generated 34,633 tissue-specific eRNA-TWAS prediction models using the GTEx and TCGA reference panels. These models covered 8,498 unique eRNAs. The number of eRNA-TWAS prediction models was highly correlated with the sample size of the reference panels (Fig. S5B). The average in-sample prediction accuracy for eRNA-TWAS models was 80.45%, similar to that for previous gene expression TWAS models. These results indicate that similar to gene expression TWAS models, most *cis*-regulated eRNA is captured by *cis*-SNPs.

To characterize the landscape of cancer susceptibility eRNAs, we applied our prediction models to 57 GWAS summary statistics of 23 cancer types. Our analysis revealed 626 susceptibility eRNAs associated with cancer susceptibility across these different cancer types (false discovery rate (FDR)<0.05; Fig. 5A; Table S5). As previously described ^10^, eRNA-linked genes were determined based on their proximity ( ≤ 1MB) and significant co-expression (Spearman’s correlation Rho ≥L0.3 and FDRL<L0.05). Consequently, we identified a set of 1,011 eRNA-linked genes that exhibited associations with the 23 distinct cancer types. Notably, 54.90% of eRNA-linked cancer susceptibility genes were specifically identified through eRNA-TWAS but not gene expression TWAS (eTWAS). For instance, within the eRNA-TWAS-identified breast cancer susceptibility eRNA-linked genes, two widely recognized breast cancer susceptibility genes, *BRCA1* and *FGFR2* (Fig. 5B), were found to have statistically significant associations (FDR < 0.05). Additionally, in the context of prostate cancer, *CCND1*, a well-established gene associated with prostate cancer susceptibility, was also identified (Fig. 5B).

**Fig. 5.**
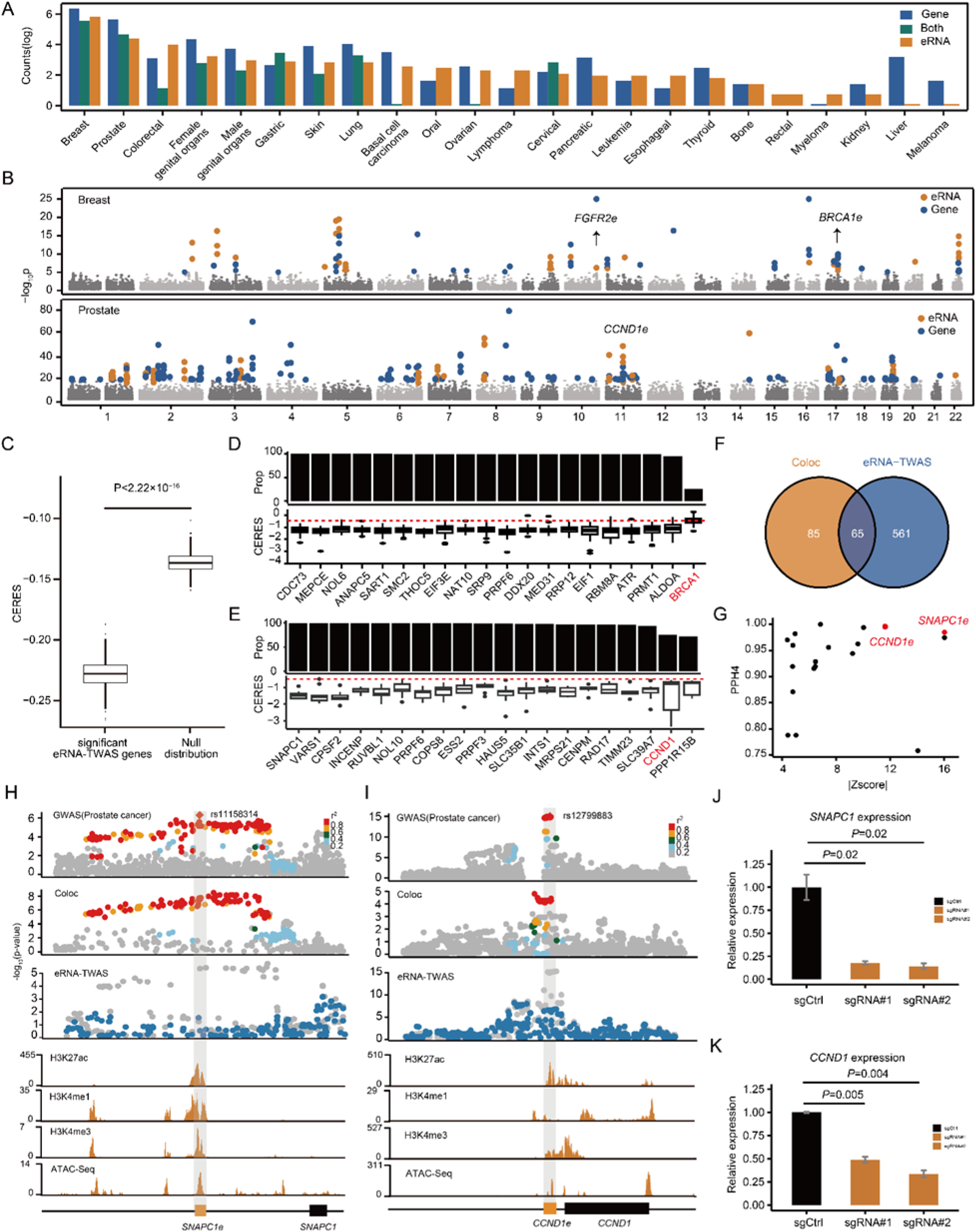
Landscape of cancer susceptibility eRNAs across 23 cancer types. (A) The eRNA- TWAS models successfully identified 626 cancer susceptibility eRNAs and 1,011 eRNA-linked cancer susceptibility genes across 23 cancer types. Notably, among the 1,011-cancer susceptibility eRNA-link genes, 54.90% were not detected by eTWAS. (B) A Manhattan plot was meticulously constructed to visually represent TWAS findings for both breast and prostate cancer. eRNAs identified via eRNA-TWAS are marked in yellow for easy visual identification, whereas genes uncovered through eTWAS are marked in blue. Each dot signifies the negative logarithm (to base 10) of the *P*-value associated with each eRNA and gene identified through eRNA-TWAS and eTWAS, respectively, plotted along the y-axis. (C) Box plots displaying data from the Sanger DepMap Project Score highlighting cancer susceptibility eRNA-link genes contributing to cell proliferation. In this context, ‘significant eRNA-TWAS genes’ refers to eRNA-linked genes identified as being associated with cancer through eRNA-TWAS. Conversely, ‘Null distribution’ refers to eRNA-linked genes unrelated to cancer. CERES scores were used to assess the essential levels of genes considering the computational effects of copy number and depletion of gene-targeting guide RNAs. Red dashes represent the median CERES score cutoff value of < -0.5, indicating a crucial role in cell proliferation. (D) Proportion of cell lines exhibiting CERES scores < -0.5 for genes associated with eRNA-linked breast cancer susceptibility (top panel) and the top 20 cancer susceptibility eRNA-linked genes with the lowest CERES scores evaluated in breast cell lines (bottom panel). *BRCA1*, a well-established breast cancer susceptibility gene, was used as a positive control. (E) Proportion of cell lines exhibiting CERES scores < -0.5 for genes associated with eRNA-linked prostate cancer susceptibility (top panel) and the top 20 cancer susceptibility eRNA-linked genes with the lowest CERES scores evaluated in prostate cell lines (bottom panel). *CCND1,* a well-known oncogene in prostate cancer, was used as a positive control. (F) Intersection between cancer susceptibility eRNAs identified through colocalization analysis and eRNA-TWAS. (G) Correlation between the absolute effect sizes (z-scores) as assessed by eRNA-TWAS and the PPs (PP_H4_) derived from colocalization analysis for significant eRNAs linked to prostate cancer susceptibility. Points highlighted in red denote eRNAs with both high PP in colocalization analysis and substantial effect sizes in eRNA-TWAS, underscoring the eRNAs that exhibit strong evidence of association with prostate cancer susceptibility. (H) LocusZoom plot illustrating the association of prostate cancer GWAS SNPs and eRNA-QTLs at the *SNAPC1* eRNA (*SNAPC1e*) locus. Notably, a concurrent association was identified between the variant rs11158314 with both the risk of prostate cancer (first panel) and the expression level of *SNAPC1e* (second panel). Manhattan plot depicting prostate cancer GWAS signals before and after conditioning on *SNAPC1e* expression (third panel). Genome browser visualization highlights the landscape of histone modifications and ATAC-seq peaks at the *SNAPC1e* locus in the PC3 cell line (bottom panels). Histone modification and ATAC-seq data were obtained from the ENCODE Project, providing a comprehensive view of the regulatory elements influencing the *SNAPC1e* locus. (I) LocusZoom plot illustrating the association of prostate cancer GWAS SNPs and eRNA-QTLs at the *CCND1* eRNA (*CCND1e*) locus. Notably, a concurrent association was identified between the variant rs12799883 with both the risk of prostate cancer (first panel) and the expression level of *CCND1e* (second panel). Manhattan plot depicting cancer GWAS signals before and after conditioning on *CCND1e* expression (third panel). Genome browser visualization highlights the landscape of histone modifications and ATAC-seq peaks at the *CCND1e* locus in the PC3 cell line (bottom panels). Histone modification and ATAC-seq data were obtained from the ENCODE Project, providing a comprehensive view of the regulatory elements influencing the *CCND1e* locus. (J) Repression of *SNAPC1e* mediated by Zim3-KRAB-dCas9 resulted in decreased expression of SNAPC1 as determined by quantitative PCR analysis in PC3 cells. This observation was statistically validated using a two-sided t-test, based on data from three independent experiments, with results presented as mean ± standard error. The term ‘sgRNA’ refers to single guide RNA. (K) Repression of *CCND1e* mediated by Zim3-KRAB-dCas9 resulted in a decreased expression of *CCND1* as determined through quantitative PCR analysis in PC3 cells.

Collectively, we successfully identified 626 cancer susceptibility eRNAs and 1,011 eRNA- linked cancer susceptibility genes across 23 cancer types. Notably, among these eRNA-linked cancer susceptibility genes, 54.90% were specifically identified through eRNA-TWAS but not traditional eTWAS.

### Cancer susceptibility eRNA-link genes are essential for cancer cell proliferation

To explore the biological and clinical relevance of cancer susceptibility eRNA-linked genes, we performed cancer hallmark enrichment analysis encompassing various key processes and characteristics of cancer development and progression. Our findings revealed that 343 cancer susceptibility eRNA-linked genes were involved in 10 well-established cancer hallmarks (Fig. S5C). These cancer susceptibility eRNA-linked genes were significantly enriched in multiple cancer hallmarks, specifically in sustaining proliferative signaling (*P* = 8.12×10^−7^; Fig. S5D), which indicated that the significant or substantial role of cancer susceptibility eRNA-linked genes were mainly involved in the cell proliferation. Therefore, we further analyzed the essentiality of cancer susceptibility eRNA-linked genes for cell proliferation using available CRISPR gene knockdown data ^40^. We evaluated essentiality based on the CERES score, accounting for factors such as copy number variations and guide RNA depletion. A lower CERES score indicates higher gene essentiality. Our analysis revealed that cancer susceptibility eRNA-linked genes exhibited significantly higher essentiality in promoting cell proliferation (Mann-Whitney U test, *P* < 2.2**×**10^-16^; Fig. 5C). Among the eRNA-linked genes identified in the relevant tissues, a substantial number demonstrated clear evidence of their essential roles in promoting cancer cell growth (CERES score < -0.5). For example, 19 breast cancer susceptibility eRNA-linked genes exhibited comparable or even higher levels of essentiality than the well-established breast cancer susceptibility gene *BRCA1* (*BRCA1*_CERESscore_ = -0.36; Fig. 5D). Similarly, in prostate cancer, 18 eRNA-linked genes associated with prostate cancer susceptibility displayed comparable or even higher levels of essentiality than the well-known prostate cancer oncogene *CCND1* (*CCND1*_CERESscore_ = -1.20; Fig. 5E). Notably, we observed a significant degree of essentiality in prostate cell lines for *SNAPC1*, a newly identified susceptibility eRNA target gene (*SNAPC1*_CERESscore_ = -1.46). We also note that *SNAPC1* was found to be essential for the proliferation of all studied cell lines (N = 1,095; Fig. 5E), whereas *CCND1* was essential for the proliferation of 75.53% of cell lines analyzed (N = 827; Fig. 5E). These findings highlight the critical involvement of these eRNA-linked genes in driving cancer cell proliferation.

### Validation of prostate cancer novel susceptibility eRNAs

To further refine the transcript-based fine-mapping of the eRNA-TWAS outcome, we identified 65 eRNAs meeting stringent criteria including significance below a threshold (*P*_Bonferroni_ <0.05) in the eRNA-TWAS analysis, and a high probability threshold (PP_H4_>0.75) in the colocalization analysis (Fig. 5F), among which 19 were found to be eRNA-linked genes specifically linked to prostate cancer susceptibility within the fine-mapped eRNA-TWAS analysis. Notably, although some of these genes have been previously reported, they were not directly identified through TWAS analysis. Base on the effect size in the eRNA-TWAS and strong colocalization with prostate cancer, we focus on the eRNAs in regulating the well-known oncogene Cyclin D1 (*CCND1*) and a novel prostate cancer susceptibility gene Small Nuclear RNA Activating Complex Polypeptide 1 (*SNAPC1*) for further experimental validation (*Zscore* = 11.64 for *CCND1* and *Zscore* = 16.04 for *SNAPC1*, PP_H4_ = 0.99 for *CCND1* and PP_H4_ = 0.97 for *SNAPC1*, Fig 5G). Furthermore, our analysis revealed that *SNAPC1e* and *CCND1e* exhibited conditionally independent patterns at the specific prostate cancer loci under investigation (Fig. 5, H to I), suggesting that eRNA-mediated risk variants account for a considerable portion of the GWAS signal within this particular genomic region.

To experimentally confirm *SNAPC1e* as the target enhancer, we employed CRISPR interference specifically targeting *SNAPC1e* in prostate cancer cells (PC3) (Fig. 5J). This CRISPR interference resulted in a remarkable 84.20% reduction in *SNAPC1* gene expression, providing strong evidence for its role as the enhancer target. Similarly, by interfering with *CCND1e*, we observed a 58.75% decrease in *CCND1* gene expression (Fig. 5K).

In conclusion, our comprehensive analysis integrating colocalization and eRNA-TWAS techniques successfully identified numerous novel cancer susceptibility eRNA-linked genes. Among them, *SNAPC1* and *CCND1* emerged as promising candidates that underwent rigorous experimental validation. The consistent identification of these genes in both the colocalization and eRNA-TWAS analyses, combined with their crucial involvement in the proliferation of prostate cancer cells, highlights their potential significance in cancer development.

## Discussion

GWAS has been highly effective in identifying thousands of SNPs associated with a wide range of complex human traits and diseases. Notably, most SNPs identified through GWAS are located within noncoding regions of the genome, particularly enhancer regions that play a crucial role in regulating gene expression. Despite these findings, the biological relationship between genetic variants and disease susceptibility has remained largely unknown. A promising strategy to bridge this gap involves identifying eQTLs as essential intermediaries linking GWAS variant signals with disease phenotypes. eQTLs may provide valuable insights into how genetic variants contribute to the development and progression of diseases, whereas they can only explain a small proportion of disease-related variants. Therefore, further research is needed to understand the molecular mechanisms contributing to complex diseases.

Here, we conducted QTL mapping for expressed eRNAs utilizing 49 normal tissues from the GTEx and 31 tumor types from the TCGA. Our analysis identified 11,757 eRNA-QTLs within the GTEx cohort and 9,316 eRNA-QTLs within the TCGA cohort. Our CRISPR-based base editing experiment confirmed that disruption of eRNA-QTLs affects the binding motifs of TFs and modifies the expression of eRNAs. eRNA-QTLs exhibited greater tissue specificity than eQTLs, consistent with a previous study ^15^ reporting that eRNAs (>30%) display higher cell-type specificity than mRNAs (<5%). The large-scale eRNA-QTL atlas allowed the generation of a genetic map between regulatory variants and complex human traits. By performing SMR and colocalization analyses, we revealed that 70.02% of colocalization events were unique to eRNA- QTLs and not shared with eQTLs. Our eRNA-TWAS analyses identified 626 eRNAs associated with cancer susceptibility across 23 cancer types. Notably, 54.90% of the target genes of these eRNAs were overlooked by traditional gene expression studies. We experimentally validated that the inhibition of two newly identified susceptibility eRNAs (*CCND1e* and *SNAPC1e*) lead to a reduction in the expression of their target genes which ultimately contributes to cell proliferation. Our findings provide additional evidence for the oncogenic potential and therapeutic liability of eRNAs. This also suggests that eRNA-QTLs can explain additional disease signals that eQTLs might miss. To gain a deeper understanding of the functional interactions and relationships among these eRNA-linked genes, we conducted an integrative network analysis and revealed that cancer susceptibility eRNA-linked genes strongly converge in shared pathways, including mitotic DNA integrity checkpoint signaling (Fig. 6; hypergeometric test, *P* = 3.92×10^-7^) and the apoptotic process (hypergeometric test, *P* = 4.86×10^-5^). Overall, these results demonstrate that eRNA-TWAS can effectively identify cancer susceptibility eRNAs and their linked genes that are essential for cancer cell proliferation across various cancer types.

**Fig 6.**
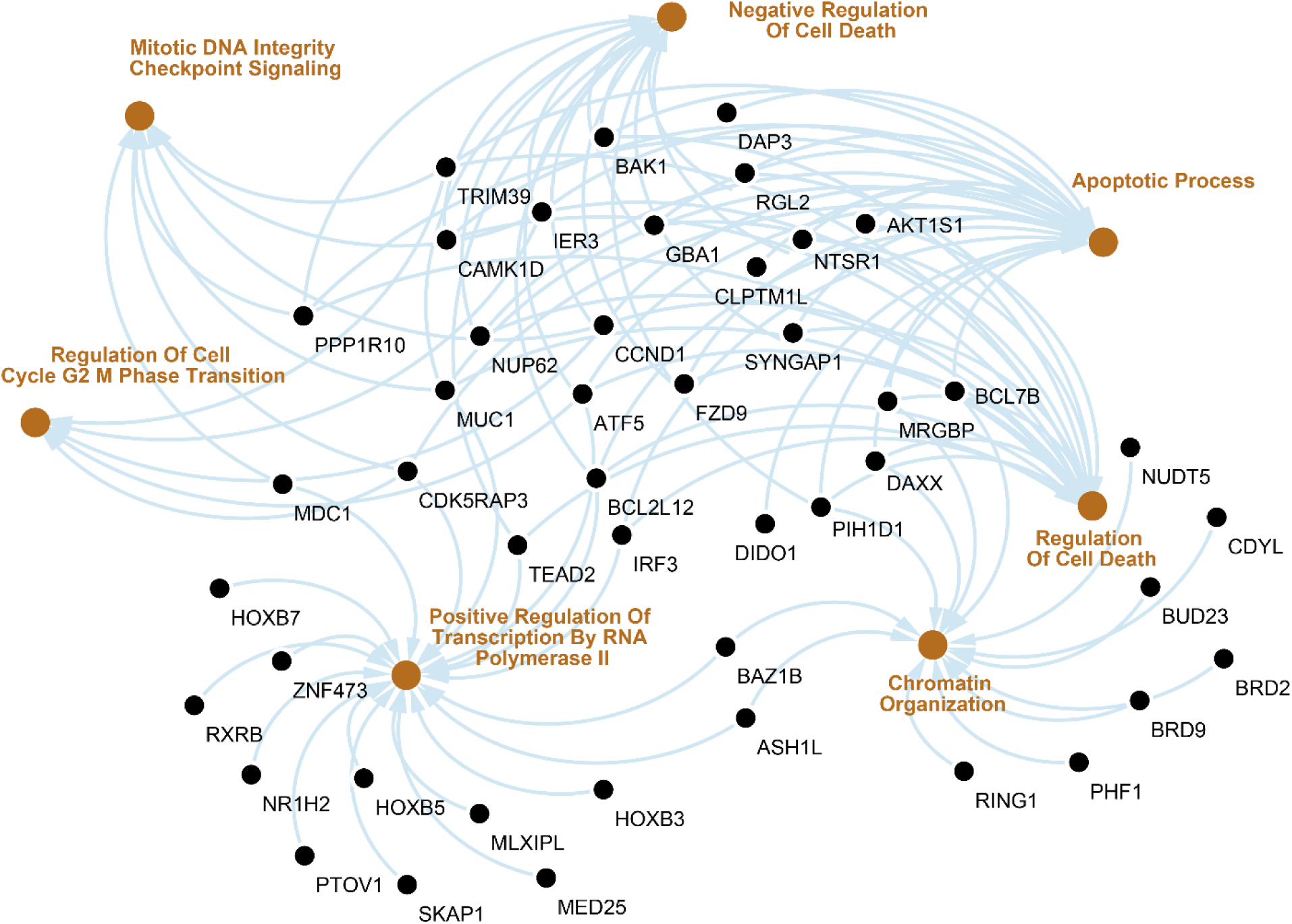
Integrative network analysis of cancer susceptibility eRNA-link genes. Integrative network analysis revealed that cancer susceptibility eRNA-link genes converge in shared pathways (yellow color), including the apoptotic process.

Notably, a single eRNA only partially contributes to target gene expression, and multiple eRNA interactions can influence target gene expression levels ^41^. For example, we discovered that the disease locus associated with inflammatory bowel disease showed colocalization with the *PSMG1* eRNA (*PSMG1e*) signal (Fig. S6A). However, this colocalization could not be explained by expression of the target gene *PSMG1*. Our SMR analysis further supports these findings, indicating that the inflammatory bowel disease locus could be explained, at least partially, by the role of *PSMG1e* (Fig. S6B). Examining single-cell level expression, we observed that *PSMG1e* was only expressed in a specific cluster, whereas its target gene *PSMG1* was expressed across all clusters (Fig. S6C). These single-cell expression data provide additional evidence of the differential expression patterns between *PSMG1e* and its target genes, highlighting the complexity of their regulatory relationships in specific cellular contexts. Considering the complexity of eRNA regulation, the eRNA-QTL strategy is a powerful approach to dissecting eRNA targets and their potential roles in disease-related processes. As demonstrated in our study, eRNA-QTL analysis provides a more comprehensive understanding of genetic regulation and disease mechanisms in cancer risk beyond what traditional eQTL analysis can capture. It expands the current strategy of molecular QTLs and supports the hypothesis that eRNA-QTLs contribute to a range of human phenotypes by regulating gene expression in a cell type-specific manner.

## Data Availability

The data presented in this study can be freely accessed, queried, visualized, and downloaded through the dedicated eRNA-QTL website portal at http://bioinfo.szbl.ac.cn/eRNA-QTL-atlas/. Sequencing data are deposited in the Gene Expression Omnibus (GEO) database (accession number: GSE242322).

## Acknowledgments

We thank members of the Li laboratory for helpful discussions. We also thank Qin Wang at Shenzhen Bay Laboratory supercomputing center for high-computing support. This work was supported by the National Natural Science Foundation of China [no. 32100533 to LL] and Open grant funds from Shenzhen Bay Laboratory [no. SZBL2021080601001 to L.L.].

## Author contributions

Conceptualization, L.L.; Methodology, W.Y.C and L.L.; Investigation, W.Y.C, J.X.L, H.C., X.X.M., Z.Y.W., and X.D.Z.; Visualization, W.Y.C., J.X.L., X.L., Y.N.W., Y.M.Q., and X.L.M.; Funding acquisition, L.L.; Supervision, L.L. and Y.B.Q.; Writing—original draft, W.Y.C.; Writing—review & editing, L.L. and Y.B.Q.

## Data and materials availability

The data presented in this study can be freely accessed, queried, visualized, and downloaded through the dedicated eRNA-QTL website portal at http://bioinfo.szbl.ac.cn/eRNA-QTL-atlas/. Sequencing data are deposited in the Gene Expression Omnibus (GEO) database (accession number: GSE242322). All data are available in the main text or supplementary materials.

## STAR Methods

### Experimental Design

The following analytical methods were carried out to (i) investigate the genetic variant effects on eRNA expression and (ii) unveil new cancer susceptibility eRNAs and genes.

### Study subjects

We employed a comprehensive dataset comprising 28,033 RNA sequencing samples from 11,606 individuals. This dataset includes 838 genotype datasets and 17,265 genotype-matched RNA-seq datasets across 49 human normal tissues sourced from the GTEx project ^23^. Additionally, we incorporated 10,768 genotype-matched RNA-seq datasets across 31 human tumor types obtained from the TCGA dataset.

### eRNA annotation

We integrated ENCODE, FANTOM, and Roadmap Epigenomics datasets to annotate enhancers. We only considered enhancers that appeared in at least two datasets. Similar to previous studies ^10^, a 3-kb region around the center of the enhancer was defined as an eRNA region. To avoid the potential interference of known transcripts, we only kept intergenic eRNA that did not overlap with available annotations, including protein-coding RNAs and ncRNAs.

### GTEx data collection and quality control

We acquired a comprehensive dataset comprising RNA-seq BAM files from 17,382 human normal samples across 54 tissues in 948 individuals obtained from the GTEx project (dbGaP, phs000424.v8.p2) ^23^. To align the original RNA-seq reads to the Human Reference Genome Build GRCh38 (hg38), we employed STAR ^42^ following the alignment parameters specified in the GTEx study ^23^. Rigorous measures were taken to ensure the integrity of the data, including the exclusion of BAM files generated from diseased tissues and tissue types with limited sample sizes. To maintain consistency and reliability, RNA-seq BAM files lacking genotype data were also removed from the analysis freeze, as they were not included in the GTEx study.

Genotype information was derived from whole-genome sequencing data obtained from the GTEx v8 release ^23^. Burrows-Wheeler alignment ^43^ was utilized to align whole-genome sequencing reads to the Human Reference Genome Build GRCh38 (hg38), and GATK HaplotypeCaller v3.5 was employed to call variants in variant call format. Subsequently, a stringent quality control process implemented by the GTEx Consortium led to the exclusion of low-quality samples, resulting in a final analysis freeze set that encompassed variants called from 838 donors. To enrich the dataset further, imputation and phasing techniques using SHAPEIT v2^44^ were applied to the final variants. Additionally, sample description files providing valuable contextual information were downloaded from the GTEx Portal (www.gtexportal.org) and associated with the analyzed samples. SNPs were excluded if they: 1) had a low call rate (< 95%), 2) had a low minor allele frequency (MAF < 1%), and 3) were out of Hardy-Weinberg equilibrium (*P* < 1×10^-6^). Samples with a low call rate (< 95%) were excluded.

### TCGA data collection, imputation, and quality control

We obtained a comprehensive dataset comprising approximately 11,000 human tumors across 31 diverse cancer types from the legacy archive of TCGA ^45^ (https://portal.gdc.cancer.gov/legacy-archive). The RNA-seq raw data underwent processing by the TCGA consortium. We utilized STAR ^42^ to align the RNA-seq data to the Human Reference Genome Build GRCh38 (hg38).

The genotype data obtained from the TCGA legacy archive were genotyped using Affymetrix Genome-Wide SNP 6.0 arrays. Birdseed genotyping files containing information on approximately 905,600 variants across a cohort of approximately 11,000 samples were downloaded for further analysis. Genotype data were aligned to the Human Reference Genome Build GRCh38 (hg38). We utilized IMPUTE2 software to impute genetic variants for the TCGA samples. The imputation process used the 1,000 Genomes Project as a reference panel. To ensure the reliability of the imputed data, we applied stringent quality control measures. Specifically, we considered only those variants with an imputation confidence score (INFO) ≥ 0.8, MAF ≥ 1%, SNP missing rate < 5%, and Hardy-Weinberg equilibrium *P* > 1 × 10^-6^. These strict criteria were employed as a cutoff to retain only high-confidence SNPs for subsequent analyses.

### eRNA quantification

Quantification of eRNA expression was conducted through eRNA annotation. We applied reads per million to quantify eRNA expression and then used the inverse-normal transformation method for normalization. We kept only eRNAs with ≥1 reads per million for subsequent analyses.

### eRNA-QTL mapping

eRNA-QTL analysis was conducted with QTLtools version 1.2 ^24^. We employed a linear regression model to adjust for covariates and used the correct mode to regress out covariates from GTEx and TCGA sample expression data. The molecular phenotype data obtained from RNA sequencing had variability from biological and technical factors. To address technical variability while preserving biological variability, we accounted for three types of covariates: 1) sex, age, body-mass index, PCR, and platform for GTEx and age, gender, tumor grade, and stage for TCGA using the metadata provided in GTEx ^23^ and TCGA ^45^, respectively; 2) the first 50 genotype principal components (PCs) derived from individuals’ genotypes to correct for population stratification observed between samples; and 3) phenotypes PCs. We performed phenotype PC analysis using QTLtools software’s ‘pca’ mode to capture experimental/technical variability by centering and scaling the expression data. To determine the optimal number of phenotype PCs capturing technical variability for eRNA-QTL discovery in GTEx and TCGA samples, we conducted multiple rounds of eRNA-QTL mapping. Each round incorporated the 50 PCs from genotypes and gradually added 0, 5, 10, 20, 30, 40, 50, 60, and 70 phenotype PCs as covariates. Through this iterative process, we selected PCs that maximized eRNA-QTL discovery in GTEx and TCGA samples.

For eRNA-QTL testing, we performed permutations with 1,000 repetitions to establish the null distribution of associations for each eRNA individually. Using the qvalue package in R ^24^, we achieved an FDR of < 0.5 for multiple tests. Moreover, we rank-normalized the quantifications on a per-phenotype basis across all samples using the –normal option in QTLtools, ensuring a normal distribution with a mean of 0 and standard deviation of 1 (N (0, 1)). Nominal *P*-values for all SNP-eRNA pairs within the cis-window (1M bp) were obtained with the nominal pass implemented in the QTLtools package. Associations between SNPs and eRNAs reaching the significance threshold corresponding to an FDR of < 0.05 were retained for further analysis.

### Conditional eRNA-QTL discovery

To evaluate the influence of additional genetic variants on the expression of a specific eRNA, we conducted a conditional analysis for each eRNA using the ‘-mapping’ option and a forward-backward stepwise regression implemented in QTLtools ^4^. The inclusion of these factors as covariates in the QTL mapping model enabled the assessment of whether an observed QTL was independent of other genetic factors.

### GWAS summary statistics curation and integration

We collected GWAS summary statistics from various sources, including published literature, the UK Biobank Imputed Dataset v.3, FinnGen Biobank, and JENGER, as these datasets were publicly accessible. Our selection of studies was based on the availability of the original publications, clear recording of population-related information, and adequate sample sizes. To avoid duplication, we identified and retained the dataset with the most comprehensive information whenever redundancy occurred across different sources. We extracted essential details such as sample size, population, and data sources from the original studies.

Our analysis focused only on GWAS data with population information precisely mapped to European ancestry individuals. To ensure data quality, we excluded individuals from consideration if their sample sizes were < 50,000, and we removed any studies that potentially had duplicated patients or controls. To assess the quality of the remaining GWAS summary statistics, we conducted a thorough examination using the R package xQTLbiolinks ^46^. This involved analyzing quantile-quantile (QQ)-plots to identify any inflation issues and P-Z plots to evaluate the consistency of analytical parameters such as beta values, standard errors, and *P*- values. Following this rigorous quality check, we identified 57 cancer GWAS summary statistic datasets deemed suitable for further downstream analyses (Table S2). To ensure consistency of the cancer GWAS summary statistics, which were based on different genome versions, we utilized CrossMap ^47^. This tool was employed to convert the GWAS coordinates to the Human Reference Genome Build GRCh38 (hg38), harmonizing the GWAS data.

### Heritability estimation

We used a restricted maximum-likelihood model implemented in GCTA ^26^ to estimate the total heritability of eRNAs arising from common genetic variants (MAF > 0.01). Heritability, in this context, refers to the proportion of phenotypic variation that can be attributed to the total genetic variation across all assessed loci. To estimate total genetic variation, GCTA generates a ‘genetic relatedness matrix’ that captures overall genetic dissimilarities among individuals in the study cohort. This comprehensive approach provides insights into the contribution of genetic factors to the observed variation in eRNA expression levels, shedding light on the heritability of these regulatory elements.

### FORGE2 enrichment analysis

We performed enrichment analysis of our eRNA-QTLs with functional annotation using the recently developed FORGE2 method ^31^ (https://forge2.altiusinstitute.org/). This method is proficient in identifying enrichment within epigenomic regions, encompassing Dnase I hotspots, histone mark broadPeaks, and hidden Markov model chromatin states. FORGE2 utilizes epigenetic data from ENCODE, BLUEPRINT, and Roadmap and evaluates the enrichment of overlap with candidate functional annotation for eRNA-QTLs compared with a matched set of background SNPs. To assess the enrichment of eNRA-QTLs across epigenomic regions, a permutation test was conducted with 1,000 iterations utilizing the regioneR package.

### Evaluation of eRNA-QTL sharing between tissues

To identify patterns of tissue sharing and tissue specificity, we performed masher analyses using a multivariate adaptive shrinkage approach implemented in the R package mashR ^27^. mashR computes posterior estimates of eRNA-QTL effect sizes and standard errors across tissues with multi-tissue eRNA-QTL summary statistics. We defined tissue sharing as an effect size within a factor of 0.5 in the same direction. Tissue specificity was described as a local false sign rate <L0.05 and z-score of at least a 2-fold difference.

### eRNA-QTL enrichment in genomic annotations

To gain insights into the functional enrichment of identified eRNA-QTLs, we performed functional enrichment analyses using torus software ^29^ following a similar approach as the GTEx Consortium. To perform the annotations, we used datasets from various sources. ENCODE provided gene regulatory elements and open chromatin annotations, and Ensembl contributed gene body annotations. Chromatin state predictions were obtained from ROADMAP, and TF binding and CpG island annotations were collected from the UCSC Genome Browser. For each specific tissue, the torus analysis generated enrichment estimates in point estimates derived using maximum-likelihood estimation. These estimates represent the logarithm of the odds ratio. To evaluate enrichment across multiple tissues, we employed a random-effects model to model the single-tissue enrichment estimates (i.e., log of odds ratio).

### Causal inference by Bayesian networks for SNP-eRNA-gene triplets

Bayesian networks (BNs) are a sophisticated type of probabilistic graphical model that leverages Bayesian inference techniques to compute probabilities. By representing conditional dependencies as edges and random variables as nodes in a directed acyclic graph, BNs aim to model not only correlations but also causal relationships among variables. In the context of genetic analysis, BNs have been utilized to gain insights into the underlying network structures that generate observed data ^48^. The joint probability density in BNs can be decomposed into marginal probability functions for individual nodes and conditional probability functions for edges, capturing the probabilistic relationships between variables. Furthermore, BNs adhere to the local Markov property, which states that each variable is conditionally independent of its non-descendants given its parent variables. For this study, we employed BNs to learn causal relationships among triplets of variables consisting of a genetic variant, eRNA, and gene. We focused on three distinct network topologies relevant to the hypotheses being tested (Fig. S3A), which included: 1) the causal model, in which the genetic variant influences the eRNA first followed by the gene; 2) the reactive model, in which the genetic variant influences the gene first followed by the eRNA; and 3) the independent model, in which the genetic variant independently influences both the gene and eRNA.

Importantly, we considered only network topologies that assumed that the signal systematically originates from the genetic variant. In practice, we applied BNs to data obtained from QTL mapping, which involved analyzing associations between eRNA-gene pairs using an approach similar to that described above. To account for multiple tests, we applied an FDR threshold of < 0.05 using the ‘p.adjust’ function in R programming language. We retained only significant results at an FDR < 0.05.

### Enrichment of eRNA-QTLs in TFBSs

To examine the enrichment of TFBSs in eRNA-QTLs and eQTLs, we analyzed ChIP-seq data for 194 TFs and 17 histone marks from the ENCODE project, constructing 2×2 contingency tables for each TF. We compared QTL variants to a null distribution of similar variants lacking regulatory associations. This null distribution was generated by sampling 1,000 random regulatory genetic variants for each QTL variant, matching them based on the relative distance to the transcription start site and minor allele frequency (within 1%). Enrichment for TFBSs was calculated as the proportion of regulatory associations within that TFBS compared with all regulatory variants relative to the same proportion in the null distribution of variants. The *P*- value for this enrichment was determined using a Fisher exact test. To account for multiple tests, we applied an FDR threshold of < 0.05 using the ‘p.adjust’ function in R programming language.

### Fine mapping of GWAS loci

Fine-mapping analysis was conducted on our curated cancer GWAS summary statistics using ancestry-matched linkage disequilibrium (LD) information. We utilized a recent toolkit that integrates three fine-mapping methods: PAINTOR (v.3.0), CAVIARBF (v.0.2.1), and FINEMAP (v.1.3.1). Each causal block was constrained to contain only one causal variant, and we applied the recommended parameters for these tools. These fine-mapping methods yield the PP of each variant as the causal one within a specified model. Subsequently, we identified credible sets consisting of variants with cumulative posterior inclusion probability (PP) values surpassing the 95% threshold.

### Enrichment of eRNA-QTLs and eQTLs within cancer GWAS risk loci

We employed fgwas (v.0.3.6) to investigate the enrichment of molecular QTLs within cancer GWAS risk loci. GWAS loci were annotated as eRNA-QTLs or eQTLs using a binary classification approach. We specifically focused on molecular QTLs that exhibited statistical significance with an FDR threshold of <0.05. Additionally, we employed quantile-quantile plots (Q-Q plots) to visualize the *P-*values of cancer GWAS SNPs. These plots provide a useful visual representation to assess deviations from the expected null distribution, aiding in the identification of potential associations. To further explore the enrichment of heritability attributed to eRNA- QTLs and eQTLs within GWAS risk loci, we applied stratified LD score regression (v1.0.1) to the cancer GWAS summary statistics. Our analysis integrated functional categories into the ‘baseline-LD model’ encompassing 53 additional functional categories. Distinct binary annotations were created for eRNA-QTLs and eQTLs, with a value of 1 assigned to the most significant eRNA-QTLs and eQTLs, whereas the remaining SNPs were assigned a value of 0. LD scores for the SNPs were computed using genotype data from individuals of European ancestry obtained from the 1,000 Genome Project (phase 3), utilizing a window size of 1 cM. Ultimately, we calculated the heritability enrichment of each category by comparing the proportion of heritability explained by the category to the proportion of SNPs within that category.

### SMR analysis

SMR analysis is a statistical method used in genetic epidemiology to investigate the causal relationship between a phenotype or trait and molecular phenotype. It involves the integration of summary-level data from GWAS and QTL studies to examine whether the effect of a genetic variant on the phenotype is mediated through molecular phenotypes. SMR analysis helps identify potential causal relationships among genetic variants, molecular phenotypes, and complex traits. Here, we conducted SMR analysis to test whether the effect of a common genetic variant on a phenotype is mediated by eRNA expression. We further performed the heterogeneity in dependent instruments (HEIDI) test to detect the existence of LD in the genetic association. *P*_HEIDI_ < 0.05 indicates that the observed genetic association could be due to LD between SNPs. The significance threshold of SMR was set at *P*_SMR_L<L0.05/N and *P*_HEIDI_L>L0.05, with N indicating the number of tests.

### Colocalization analyses

We employed a Bayesian method for colocalization analysis to determine whether there is shared causal genetic variation between molecular traits (e.g., eRNA expression) and a disease trait. Specifically, we used the R package ‘coloc’ ^38^ to assess the colocalization of cancer GWAS summary statistics and eRNA-QTL and eQTL signals. The coloc package calculates five hypotheses: H0 (no association), H1 (GWAS association only), H2 (eRNA-QTL or eQTL association only), H3 (both associations but not colocalized), and H4 (both associations and colocalized). We performed separate analyses for each cancer risk phenotype and each proximal eRNA or gene using default parameters. To determine whether an eRNA or gene and GWAS signal were colocalized, we set the threshold of the PP_H4_ to be > 75%. Additionally, we required the ratio of PP_H4_ to the sum of PP_H3_ and PP_H4_ (PP_H4_/(PP_H3_ + PP_H4_)) to be ≥ 0.9. These criteria helped identify cases where there was strong evidence for both the eRNA or gene and GWAS associations being driven by the same underlying causal variants. We employed LocusZoom (v.1.4) to visualize regional plots and PLINK (v.1.90) to assess the LD between the identified causal SNP and other SNPs.

### eRNA-TWAS for cancer GWAS

We utilized the FUSION framework to perform eRNA-TWAS. Our approach began by employing a mixed-linear model to estimate the heritability of the eRNA region. This estimation was based on SNPs with a MAF > 0.01 located within 1 Mb of the eRNA region using a reference panel that consisted of cohorts with matched RNA-seq and genotype data. To ensure robust covariate adjustment, we incorporated well-established factors utilized in the QTL mapping section to determine eRNA expression. Subsequently, only eRNAs with significant heritability estimates (*cis*-h^2^) below a Bonferroni-corrected *P*-value of 0.05 were retained for further analysis. Within the FUSION framework, we selected three different models for weight calculation: BLUP, ENET, and LASSO. A cross-validation approach was employed to determine the model with the optimal eRNA-TWAS prediction accuracy for each gene. Subsequently, we applied these eRNA-TWAS prediction models to GWAS summary statistics using an FDR threshold of 0.05.

### The identification of eRNA-link gene

We obtained gene annotations from ENSEMBL (https://jul2023.archive.ensembl.org/index.html), GENCODE (https://www.gencodegenes.org/human/release_38.html), and UCSC (https://genome.ucsc.edu/index.html) and integrated them. The expression matrix of these genes across human tissues was collected from the GTEx portal, and across cancer types from TCGA. Putative target genes of eRNAs were identified by assessing proximity (≤ 1MB) and co-expression (Spearman’s correlation coefficient RsL≥L0.8 and FDRL<L0.05) between individual eRNAs and their target genes across various tissues and cancer types. To ensure accuracy, eRNAs located within intronic regions of the target genes were excluded from the correlation analysis.

### Annotation of cancer susceptibility eRNA-link genes in cancer-relevant gene databases

To ascertain the intersection between our cancer susceptibility eRNA-link genes and established cancer-related genes, we compiled cancer-related gene sets from reputable sources, including the Molecular Signatures Database (MSigDB), DORGE, and Catalogue of Somatic Mutations in Cancer (COSMIC) Gene Census (https://cancer.sanger.ac.uk/census). We identified putative cancer-related genes by annotating with specific key phrases, such as ‘breast cancer’ and ‘prostate cancer’.

### Gene set enrichment analysis

We selected pan-cancer identified cancer susceptibility eRNA-link genes, filtering for those that encode HLA genes within MHC regions. We performed gene set enrichment analysis (GSEA) between the two groups to identify significantly altered Gene Ontology (GO) pathways utilizing GSEA software (v4.3.2) ^49^. GSEA is a widely utilized software package that utilizes gene sets to discern different biological functions between two groups. GO analysis encompasses biological processes, cellular components, and molecular functions. The statistical significance of enrichment was determined using the hypergeometric distribution. *P-*values generated by hypergeometric tests were FDR-corrected for multiple testing, and a *P* <L0.05 was considered statistically significant. The integrative network graphs were visualized using Cytoscape, a software tool for visualizing and analyzing biological networks.

### Cell line maintenance and generation

HEK293T cells were cultivated in Dulbecco’s Modified Eagle Medium (Gibco, cat #: C11995500BT) supplemented with 1% penicillin/streptomycin and 10% fetal bovine serum (FBS; Gibco, cat #: C10010500BT), 100 IU/mL penicillin, and 100 μg/mL streptomycin (Gibco, cat #: 15140-122) and were incubated in 5% CO_2_ at 37L. HEK293T cells were seeded onto 6- well plates and transfected with BE4max-NG or SpRY-BE4max and sgRNA plasmids using EZtrans (Shanghai Life iLab Biotech, cat #: AC04L099) according to the manufacturer’s instructions. Synthesized oligos (Table S6) were annealed and ligated into the Bsa1-digested pGL3-U6-pGK-puro vector. To screen cells expressing pGL3-U6-sgRNA-pGK-puro, puromycin (2.5 µg/ml; Merck, cat #: 540411) was added to cells after transfection for 24 h and then collected at 48 h post-treatment. Transfected cells with high mutation efficiency were seeded onto a 96-well plate with about 60 cells. After 12 days of cultivation, single clones were subjected to DNA extraction and genotyping. Clones with expected mutations were passaged for subsequent experiments.

PC-3 cells obtained from the Cell Resource Center of Shanghai Institutes for Biological Sciences (Chinese Academy Science, Shanghai, China) were cultured in RPMI-1640 (Gibco, cat #: C22400500BT) supplemented with 10% FBS, 100 IU/mL penicillin, and 100 μg/mL streptomycin. Lentivirus was produced by cotransfecting HEK293T cells with transfer plasmids and standard packaging vectors pMD2.G and psPAX2 using PEI according to the manufacturer’s instructions. To generate PC-3 cells stably expressing CRISPRi effectors, PC-3 cells were infected with lentivirus containing Zim3-KRAB-dCas9 (pLX303) at low multiplicity of infection followed by 3 days puromycin selection.

### Genomic DNA extraction and genotyping

Transfected HEK293T cells were treated with lysis buffer consisting of 1.5 mM MgCl2, 10 mM Tris-HCl (pH 8.0), 50 mM KCl, 0.5% Tween-20, 0.5% Nonidet P-40, and 100 μg/ml proteinase K (ThermoFisher Scientific). DNA fragments containing the targeting sites amplified by PCR with Phanta Max Super-Fidelity DNA polymerase (Vazyme; P505) were subjected to Sanger sequencing for genotyping. The primers used for PCR are listed in Table S6.

### Assessing transcriptional effects via targeted enhancer repression

Two guide RNA oligos per enhancer were inserted into the pBA900 (pU6-sgRNA EF1Alpha- puro-T2A-BFP) at the Blp1 and BstX1 restriction sites. gRNA constructs and control vector were separately packaged into lentivirus, which was subsequently transduced into PC-3 cells stably expressing Zim3-KRAB-dCas9. Ten days post-infection, transduced cells were sorted via flow cytometry using a BD FACS Aria3. Total RNA was extracted using the Quick-RNA™ Miniprep Kit (cat#: R1055; Zymo Research), and cDNA was generated using the Hifair® L 1st Strand cDNA Synthesis Super Mix for qPCR (gDNA digester plus) kit (Yeasen, cat #: 11141ES60). Quantitative PCR was performed using the Hieff® qPCR SYBR Green Master Mix (Yeasen, cat #: 11203ES08) on a CFX96 machine (BIO-RAD, Hercules, CA, USA). Experiments measuring the expression of each gene were repeated at least three times, with GAPDH used as the internal reference for expression. The gRNA oligos and primers used for qPCR are listed in Table S6.

### RNA-seq and analysis

Total RNA extracted from HEK293T cells was subjected to strand-specific RNA sequencing using the Illumina nova seq 6000 platform at Berry Genomics Co, Ltd (Beijing, China). Briefly, strand-specific RNA-seq libraries were prepared by combining the Ribo-Zero rRNA Removal Kit (Epicentre, Madison, WI, USA) and dUTP method to ensure strand specificity. RNA samples, namely, four samples from HEK293T cell lines under two conditions (control: wild-type; case: heterogenous) with two biological replicates per condition, were obtained in FASTQ format. For each FASTQ file, quality checks were conducted using FastQC ^50^. Contaminating data, such as low-quality reads, adaptor sequences, and poor-quality bases, were removed with Trimmomatic software ^51^. Trimmed reads were mapped to the human reference genome (GRCh38) using STAR ^42^ and then sorted by SAMtools ^52^.

### Statistical analysis

All results are presented as the mean ± 95% confidence interval. Statistical analysis was performed using R Studio (version 4.1.0). Differences between two groups were analyzed using unpaired two-tailed Student’s t-tests or Wilcoxon signed-rank tests. For multiple comparisons, one-way analysis of variance (ANOVA) with post hoc Tukey HSD tests were used. *P*-values < 0.05 were considered statistically significant. \**P* < 0.05, \*\**P* < 0.01, \*\*\**P* < 0.001, and ****P < 0.0001.

### eRNA-QTL atlas for exploration and analysis of genetic effects on eRNA

To enhance accessibility to eRNA-QTL information, we developed an intuitive and user-friendly database (eRNA-QTL atlas, http://bioinfo.szbl.ac.cn/eRNA-QTL-atlas/). We implemented the eRNA-QTL search and query functionality within our robust database infrastructure, enabling users to conduct searches across 49 distinct human tissues. This advanced capability allows users to thoroughly explore the functional specific characteristics of eRNA-QTLs by eRNA IDs (e.g., ENSR00000069687), thereby identifying associations with eRNA-QTLs of particular interest (Fig. S7A). Furthermore, direct retrieval of eRNA-QTLs is also included, which provides researchers with a sophisticated tool for their investigations. We then employed a genome browser to visualize and explore the eRNA-QTL genomic landscape surrounding target eRNAs by their genome positions or adjacent genes of target eRNAs. For example, eRNA-QTLs in a specific tissue or all tissues for ENSR00000253886 can be shown by searching for its adjacent gene *TBX15*, which is positioned upstream of the adjacent region of ENSR00000253886 (Fig. S7B). These browsers provide an intuitive interface for navigating genomic regions and visualizing annotations, including gene structures, GWAS catalog risk SNPs ^53^, and eRNA-QTLs among all 49 human tissues. By integrating the retrieved eRNA-QTL data and genome browser visualizations, we construct a comprehensive profile of the target eRNAs, shedding light on their potential function. In addition to these features, we offer the ‘GWAS-eRNA-QTLs Colocalization Event Visualization’ section by R package LocusComparer ^54^. This feature helps researchers conveniently visualize colocalization events between GWAS data and eRNA-QTLs. An illustrative instance of the colocalization plot depicts the region surrounding enhancer ENSR00000089936, along with GWAS and eRNA-QTLs *P*-values, specifically in nerve tibialb tissue (Fig. S7C). Furthermore, a compilation of eRNA-QTLs associated with GWAS allows users to delve deeper into the underlying mechanisms of eRNA-QTLs in association with human traits and diseases. This compilation includes full information on 1,625 traits/diseases from the GWAS catalog across all 49 tissues (Fig. S7D). As an example, we present an instance of eRNA- QTLs associated with the trait of telomere length in adipose subcutaneous tissue. Finally, the download page allows users to download all eRNA-QTL results across 49 human tissues for further custom analysis (Fig. S7E). A comprehensive help page provides guidance for the effective utilization of these functionalities with a series of detailed, step-by-step instructions (Fig. S7F). Altogether, using the eRNA-QTL atlas, researchers and investigators can efficiently access and analyze eRNA-QTL data, empowering them to make informed decisions and gain valuable insights into the regulatory mechanisms underlying their studies related to eRNAs. Thus, our platform provides a powerful and scientifically rigorous experience for the exploration of eRNA-QTL information.

