## Supplementary material for "Enhancer RNA transcriptome-wide association study reveals an atlas of pan-cancer susceptibility eRNAs": Fig. S

Wenyan Chen *et al.*

**This PDF file includes:**

Figs. S1 to S7

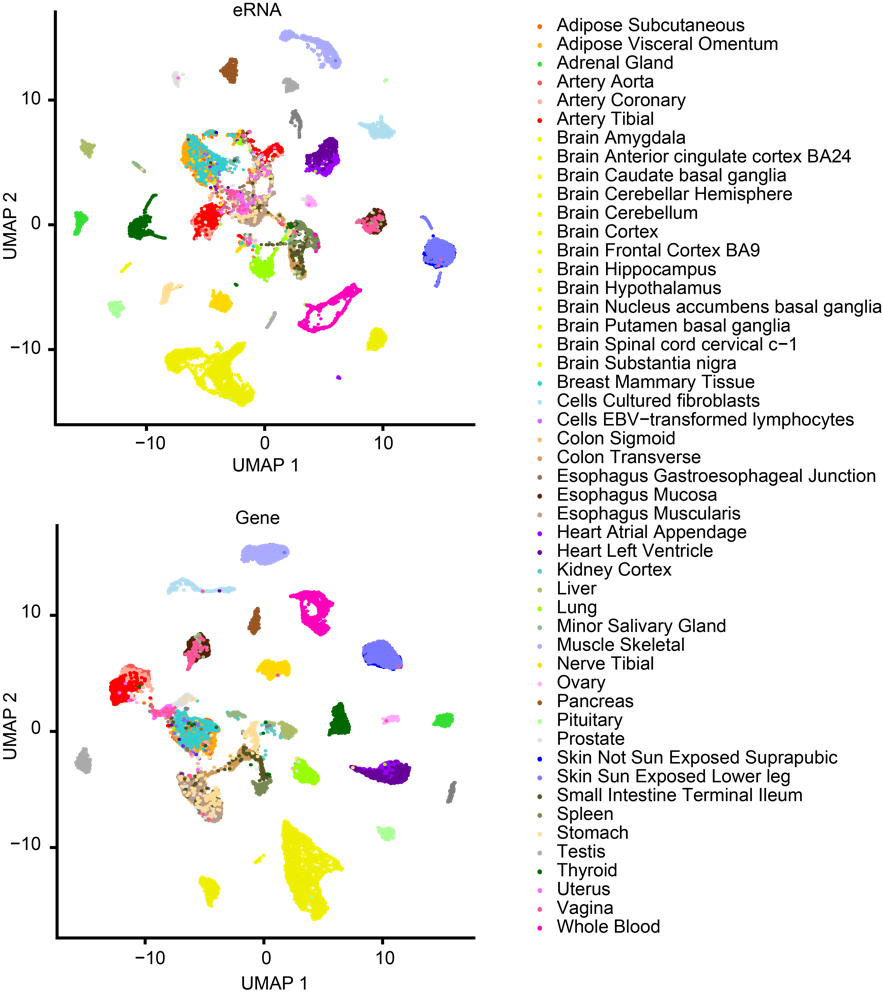

Fig. S1. UMAP of expression colored according to tissue. UMAP plot showing clustering of eRNAs and genes based on their expression in 17382 samples across 49 tissues. The color and shape of each dot refers to the tissue recorded in the GTEx dataset.

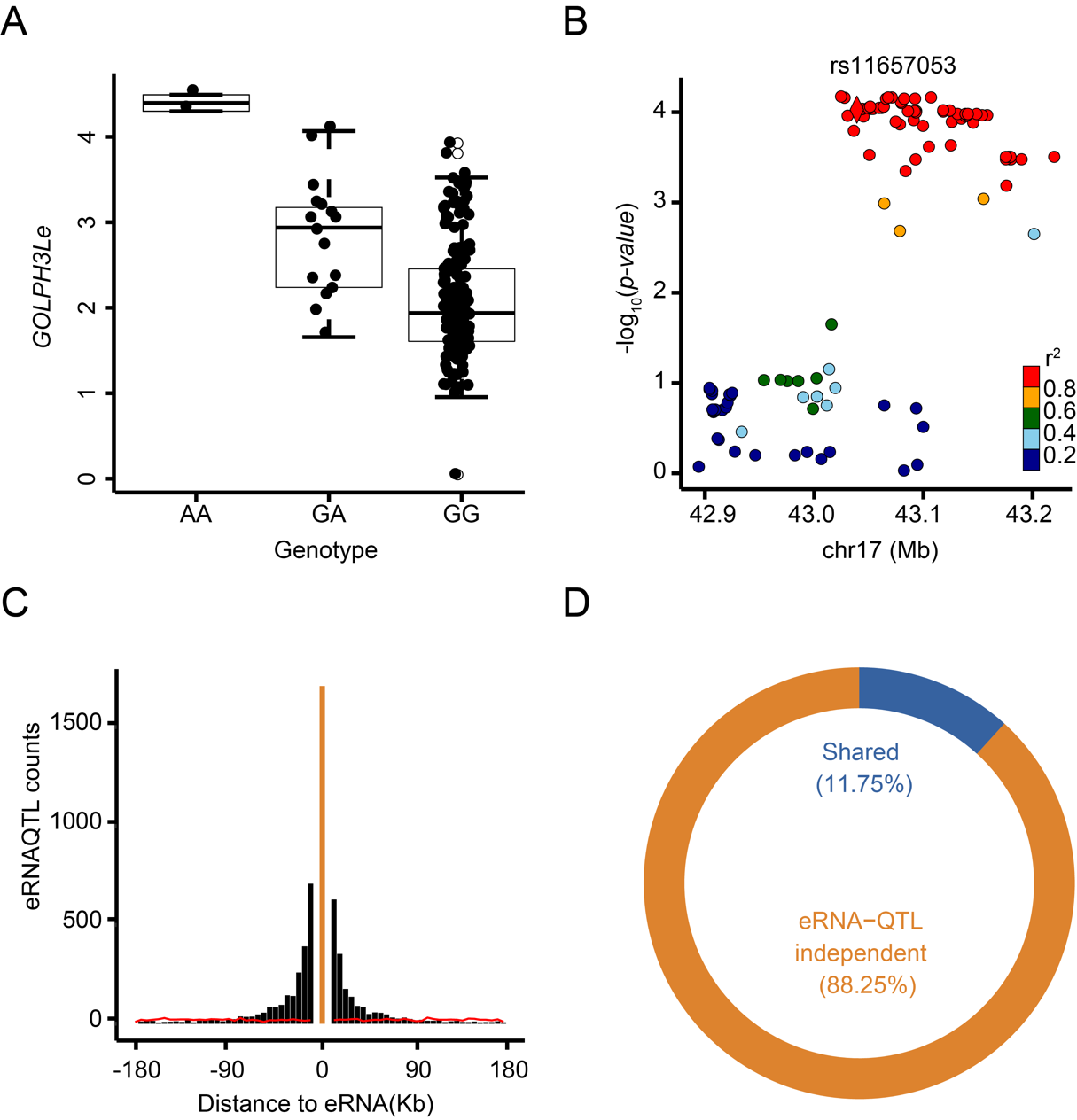

Fig. S2. **Functional Annotation of the eRNA-QTL**. (**A**) The example of SNP rs72700813 is closely related to the *GOLPH3L* eRNA (*GOLPH3Le*) in the brain cortex. Each dot in the box plot represents the expression (RPM) of *GOLPH3Le* in a particular individual (n = 184). Horizontal lines indicate the median values, and The boxes span the interquartile range between the 25th and 75th percentiles. The whiskers extend to 1.5× interquartile range (IQR). (B) *BRCA1* eRNA (*BRCA1e*), located upstream of the *BRCA1* gene, contains an independent eRNA-QTL rs11650272. (**C**) Locations of the lead eRNA-QTLs for eRNAs. The x-axis represents a candidate region divided into multiple bins, as described. The y-axis indicates the count of lead eRNA-QTLs within each bin, where each lead eRNA-QTL corresponds to a specific eRNA and represents the most significant eRNA-QTL. For eRNA-QTLs that fall outside eRNAs, they are assigned to bins based on their physical distance from the eRNAs. To provide a comparison, the red line represents randomly selected positions within a ±1-Mb window surrounding each eRNA. The yellow bar in the figure represents the eRNA region. (**D**) Overlap between eRNA-QTL and QTL.

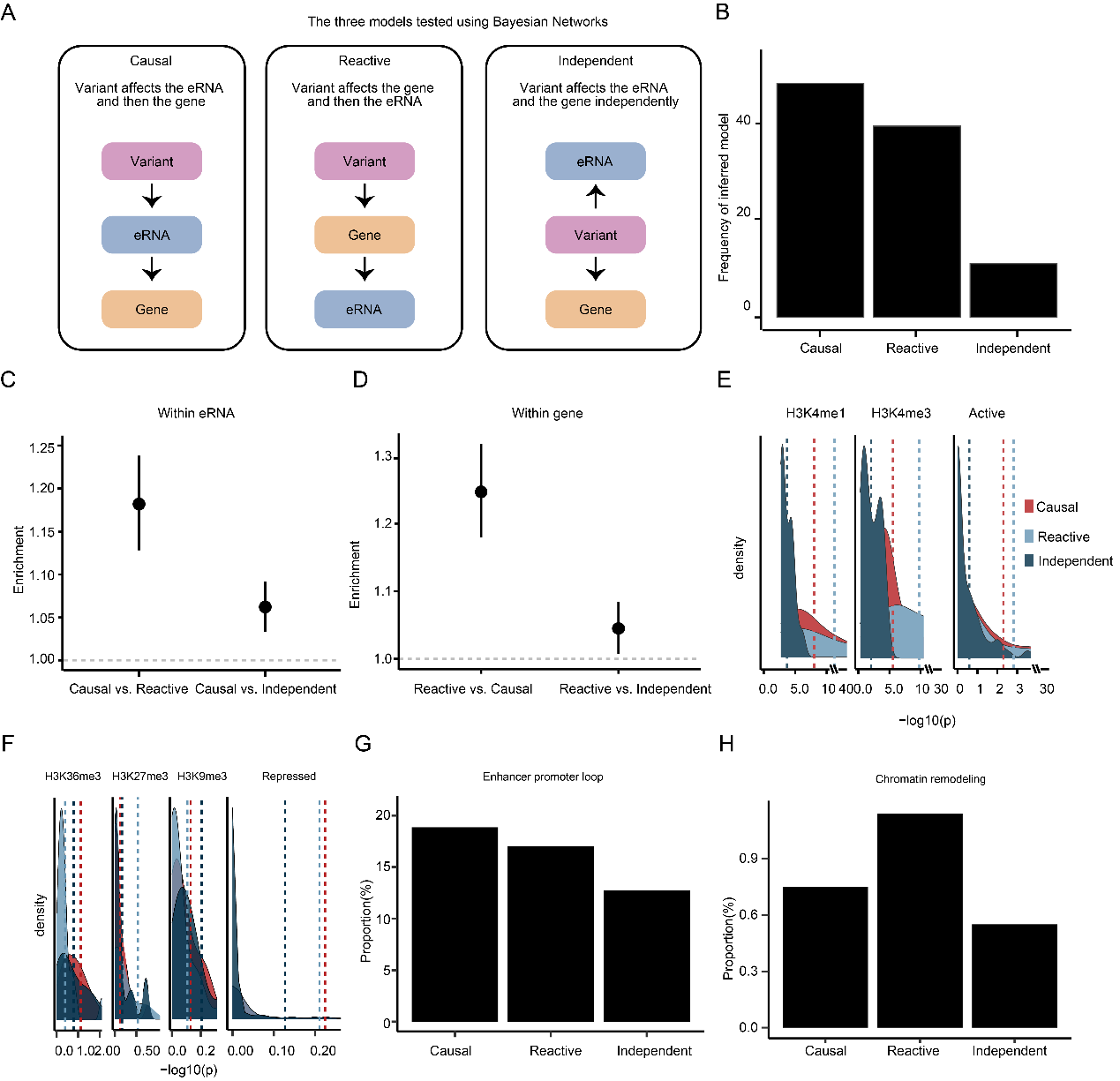

Fig. S3. eRNA-QTLs mainly affects gene expression through regulating eRNA expression. (**A**) The three models tested using Bayesian Networks. (**B**) Frequency of inferred causal model among eRNA-QTLs associated with gene expression. (**C**) Relative distances between three models (independent model, reactive model, and causal model) and their associated eRNAs/genes (**D**). (**E**) The enrichments of three models for histone mark broadPeaks and the active epigenomic marks from consolidated Roadmap Epigenomics consortium data. The dashed line represents the average p-value, which was used to assess the degree of enrichment for the three models across different histone marks and epigenomic marks. (**F**) The enrichments of three models for histone mark broadPeaks and the repressed epigenomic marks from consolidated Roadmap Epigenomics consortium data. (**G**) The proportion of genes involved in the enhancer-promoter loop was analyzed in three different models. (**H**) The proportion of genes involved in the chromatin remodeling process was analyzed in three different models.

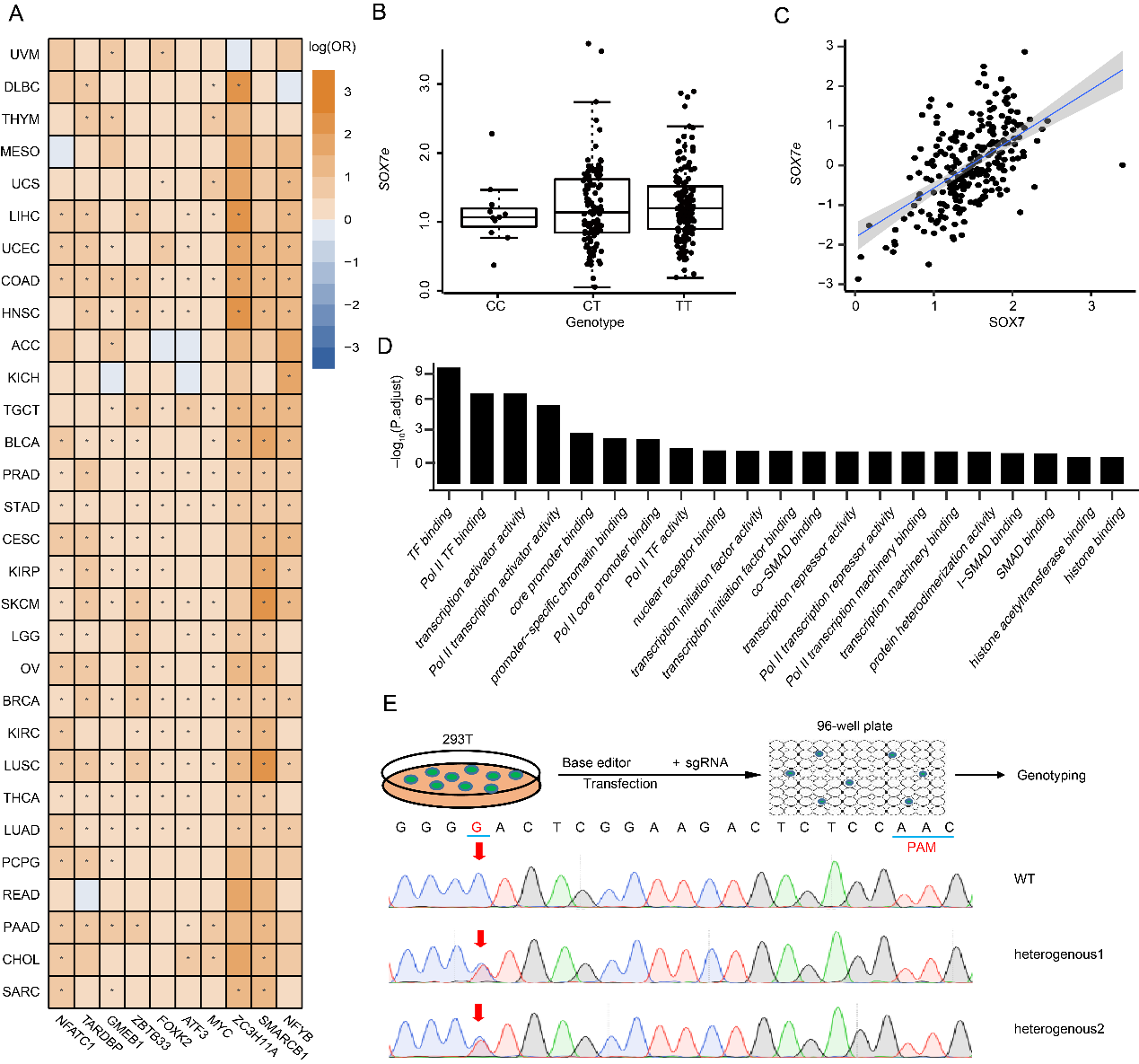

Fig. S4. eRNA-QTLs could alter transcription factor binding sites (TFBSs). (**A**) The enrichment of eRNA-QTL within the top ten TFs across a range of tumor tissues is observed. The enrichment values correspond to the maximum-likelihood estimated log (OR), and the whiskers on the plot indicate values that are below the significant false discovery rate (FDR) threshold of 0.05. (**B**) The example of SNP rs17776622 that contained the YY1-binding site is closely related to the expression of *SOX7* eRNA (*SOX7e*). (C) The expression of *SOX7e* is tightly associated with *SOX7*. (D) Biological process for TFs whose eRNA-QTL were enriched. (E) Schematic diagram of the establishment of the monoclonal cell line with expected eRNA point mutations. The eRNA-mutant bulk cells were diluted and seeded onto a 96-well plate for culture. Single-cell clones were genotyped by Sanger sequencing after culture for 12 days. The site of rs6703982 was edited by BE4max-NG.

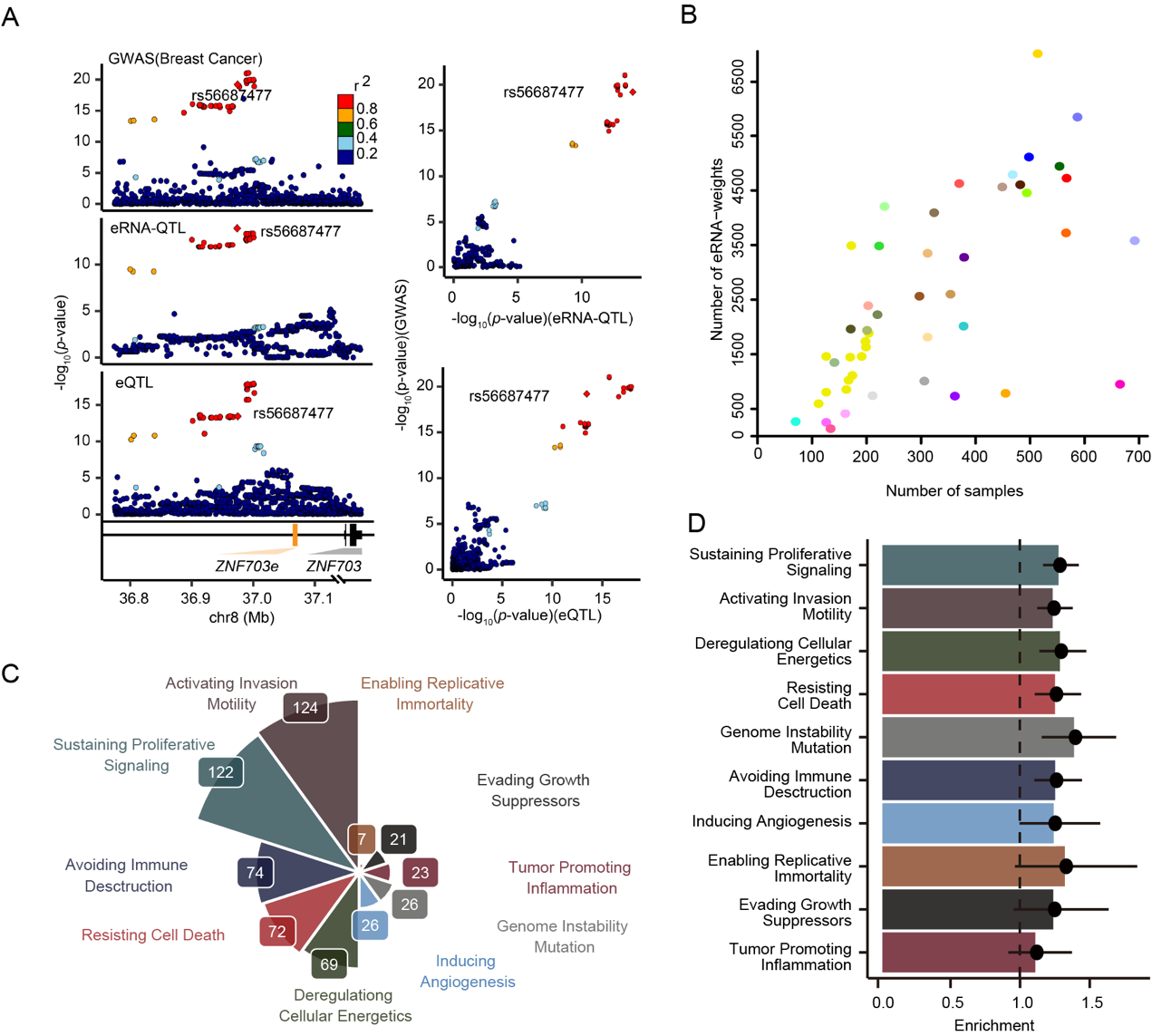

Fig. S5. The biological and clinical features of cancer susceptibility eRNA-link genes. (**A**) Shared eRNA-QTL/eQTL depict variants with regulation both on *ZNF703* eRNA (*ZNF703e*) and *ZNF703* mRNA level displayed by the significant SNP-eRNA pair rs56687477- *ZNF703e*. (**B**) The number of eRNA-TWAS prediction models was highly correlated with the sample sizes of the reference panels. The color of each dot refers to the tissue recorded in the GTEx dataset and TCGA dataset. (C) 343 cancer susceptibility eRNA-link genes are known to contribute to the set of cancer hallmarks. (D) Cancer susceptibility eRNA-link genes are enriched in the hallmarks of cancer, specifically in sustaining proliferative signaling and activating invasion motility.

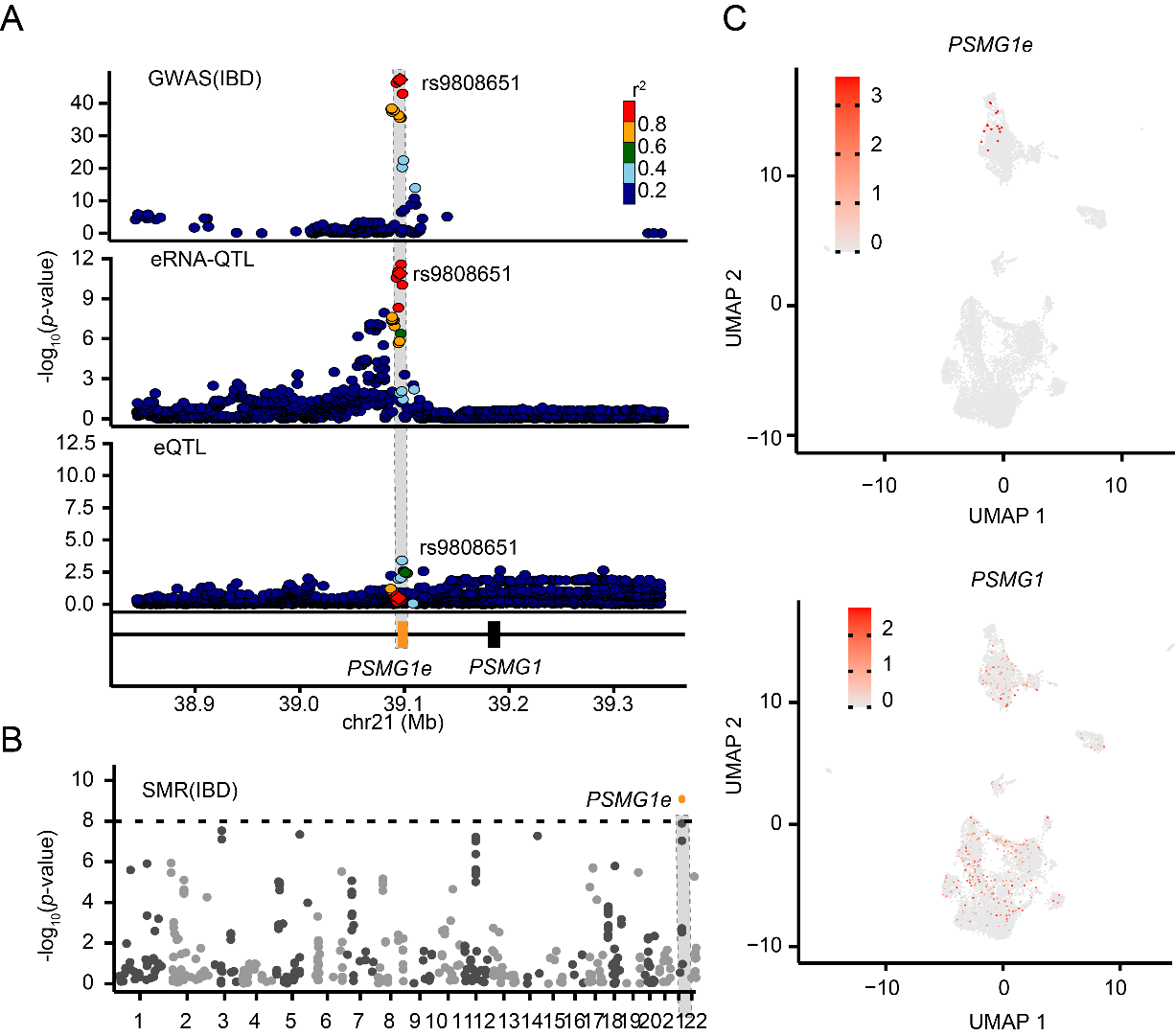

Fig. S6. **eRNA-QTLs contribute to a range of human phenotypes by regulating gene expression in a cell type-specific manner.** (**A**) locus zoom plot of inflammatory bowel disease (IBD) GWAS signal; Shading of the points represents the linkage disequilibrium (r2, based on the 1000 Genomes Project Europeans; gray indicates unknown LD) between each SNP and the top SNP rs9808651, indicated by purple shading. Second panel, locusZoom plot showing the association with *PSMG1* eRNA (*PSMG1e*) expression. Bottom panel, locusZoom plot showing the association with *PSMG1* expression. (**B**) The Manhattan plot displays the SMR result of eRNAs relevant to IBD disesase. The dashes represent the significant threshold. (C) For the visualization at the single cell level, expression UMAP plots for *PSMG1e* and the *PSMG1* were generated. Colour bar indicates log2 normalized expression.

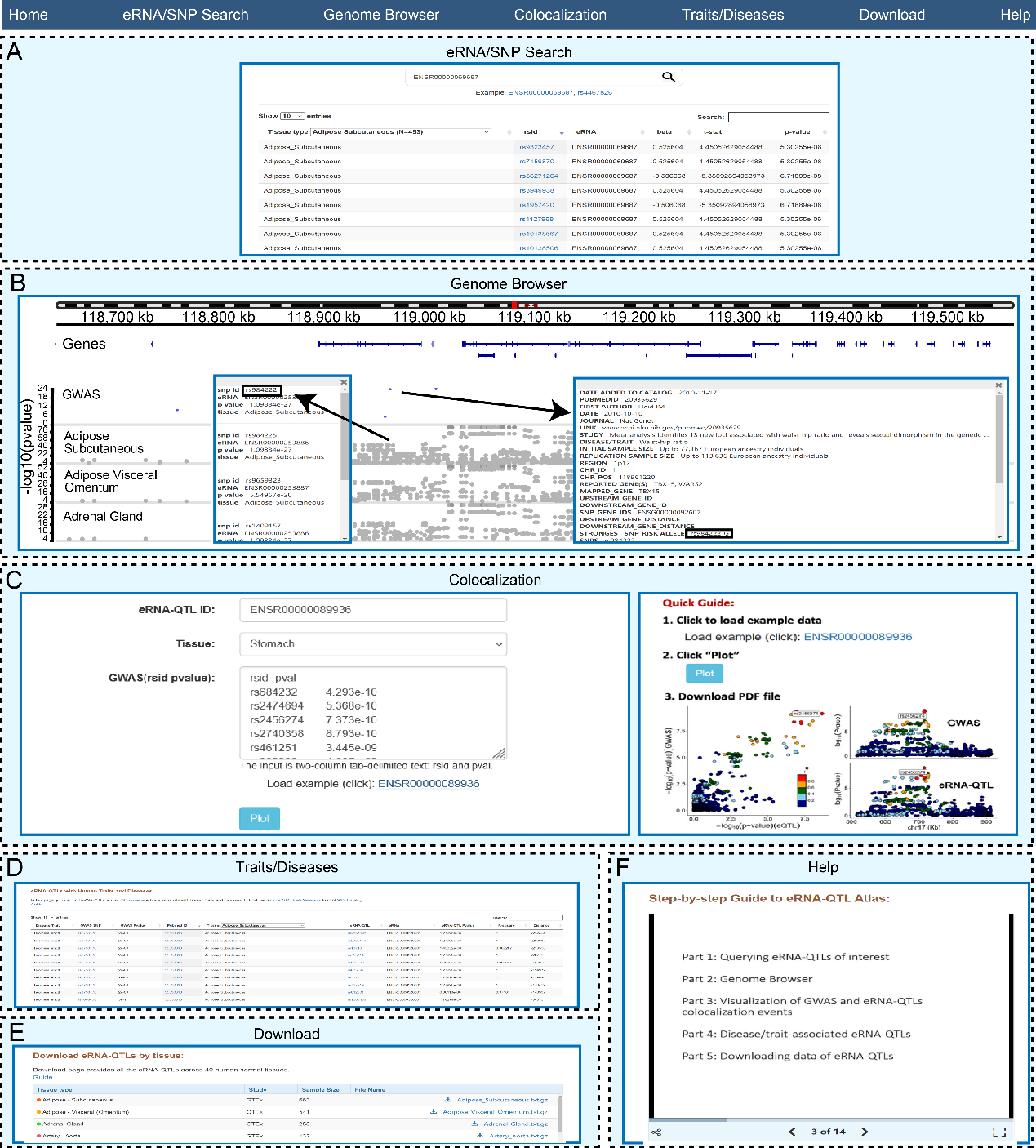

Fig. S7. **The web interface of eRNA-QTL atlas.** (**A**) eRNA-QTLs query interface and result visualization. (**B**) An example of the genome browser view shows the eRNA-QTLs in adipose subcutaneous tissue for ENSR00000253886 at the upstream of adjacent gene *TBX15*. (**C**) The interface of the ‘GWAS eRNA-QTLs Colocalization’ and an example of the LocusCompare plot at the region of enhancer ENSR00000089936 with GWAS *p-value* and eRNA-QTL *p-value* in stomach tissue. (**D**) The interface of the ‘Traits/Diseases’ and an example of the eRNA-QTLs with telomere length in adipose subcutaneous tissue. (**E**) Data download. (**F**) A detailed help page is provided to serve as a guide for users in understanding the usage of eRNA-QTL Atlas and interpreting the output data.
